# Impact of the COVID-19 pandemic on the circulation of other pathogens in England

**DOI:** 10.1101/2022.10.21.22281366

**Authors:** Lauren Hayes, Hannah Uri, Denisa Bojkova, Jindrich Cinatl, Mark N. Wass, Martin Michaelis

## Abstract

The COVID-19 pandemic and the associated prevention measures did not only impact on the transmission of COVID-19 but also on the spread of other infectious diseases in an unprecedented natural experiment. Here, we analysed the transmission patterns of 22 different infectious diseases during the COVID-19 pandemic in England. Our results show that the COVID-19 prevention measures generally reduced the spread of pathogens that are transmitted via the air and the faecal-oral route. Moreover, the COVID-19 prevention measures resulted in the sustained suppression of vaccine-preventable infectious diseases also after the removal of restrictions, while non-vaccine preventable diseases displayed a rapid rebound. Despite concerns that a lack of exposure to common pathogens may affect population immunity and result in large outbreaks by various pathogens post-COVID-19, only four of the 22 investigated diseases and disease groups displayed higher post-than pre-pandemic levels without an obvious causative relationship. Notably, this included chickenpox for which an effective vaccine is available but not used in the UK, which provides strong evidence supporting the inclusion of the chickenpox vaccination into the routine vaccination schedule in the UK. In conclusion, our findings provide unique, novel insights into the impact of non-pharmaceutical interventions on the spread of a broad range of infectious diseases.

---

Previous studies have suggested that non-pharmaceutical interventions during the COVID-19 pandemic have also affected the spread of other pathogens [1-4]. Here, we analysed the transmission patterns of 22 infectious diseases in England in the context of the COVID-19 prevention measures, using data derived from the UK Health Security Agency, the UK Office for National Statistics, and the Royal Collage of General Practitioners Research and Surveillance Centre (Suppl. Methods, Suppl. Table 1, Suppl. Table 2).

Reported cases for all investigated infectious diseases dipped in response to the first lockdown except from methicillin-resistant *Staphylococcus aureus* (MRSA), Lyme disease, and hepatitis E (Figure 1, Suppl. Figures 1-22). MRSA infections are usually diagnosed in healthcare settings [5], and some studies reported an increase of MRSA cases during COVID-19 [5]. Therefore, this finding does not seem to be surprising.

**Figure 1.**
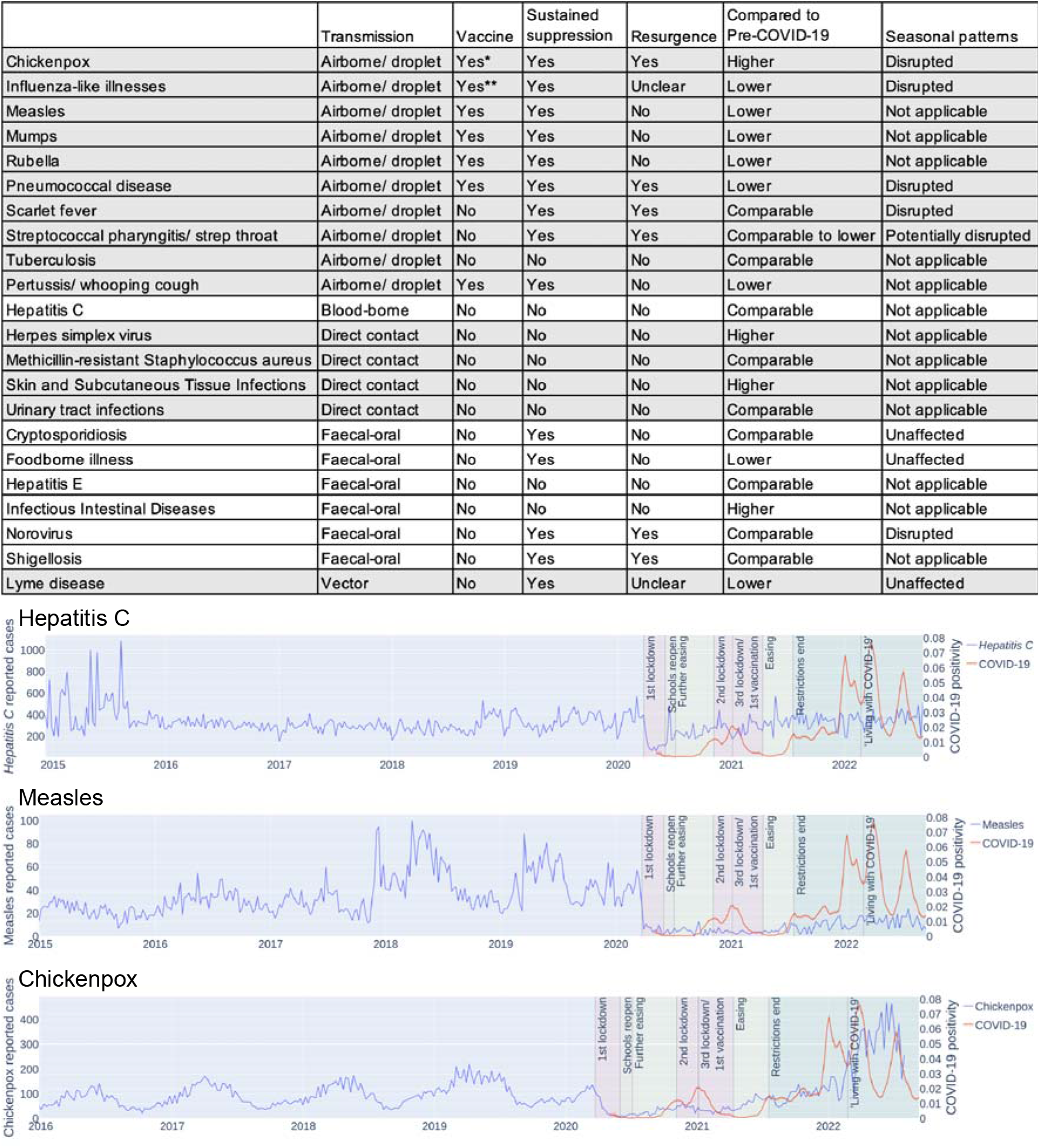
Impact of COVID-19 prevention measures on the circulation of other infectious diseases. Overview table providing a qualitative description of the impact of the COVID-19 measures on the investigated pathogens in England and curves illustrating the impact of the COVID-19 measures on hepatitis C, measles, and chickenpox. Detailed information is presented in the Suppl. Figures 1-22.

For Lyme disease, no reduction is seen in response to the initial lockdown but lower case numbers have been reported during the COVID-19 pandemic (Figure 1, Suppl. Figure 22), which is in line with other studies and commonly attributed to underreporting [6,7]. Generally, the initial drop in documented cases during the first lockdown is difficult to interpret, as it might be the consequence of underreporting [6-8].

Thirteen diseases displayed a sustained reduction during the time period when prevention measures were in place (Figure 1, Suppl. Figures 1-22). This included nine of the ten diseases that spread via the air and four of the six diseases that are characterised by faecal-oral transmission (Figure 1, Suppl. Figures 1-10 and 16-21).

The impact of the COVID-19 prevention measures on pathogens that are transmitted via the air is in agreement with other findings [3,9]. The only exception was tuberculosis (Figure 1, Suppl. Figure 9). However, most tuberculosis infections are asymptomatic and go undiagnosed [10,11]. During COVID-19, delayed diagnoses due to limited access to tuberculosis services have been suggested to have resulted in a rise of severe cases, including detrimental COVID-19/ tuberculosis co-infections [11,12]. Hence, it is plausible that the pandemic measures did not cause a reduction of severe tuberculosis cases, which are typically diagnosed.

Moreover, our findings are in line with others showing that hygiene measures and physical distancing reduce the transmission of (foodborne) enteric diseases that are transmitted via the faecal-oral route [3,8,9,13,14]. Also in agreement with previous findings [3], the COVID-19 pandemic and the related prevention measures disrupted the seasonal transmission patterns of different infectious diseases (Figure 1; Suppl. Figures 1,2,6,7,20).

There are concerns that the disruption of routine vaccinations may result in a decreased population immunity and in turn larger outbreaks of vaccine-preventable infectious diseases [3]. However, our findings indicate a sustainable suppression of vaccine-preventable diseases also beyond the lifting of restrictions (Figure 1). This included measles, mumps, rubella, pertussis, pneumococcal disease, and influenza, as indicated by the number of influenza-like illnesses (although this category may include other respiratory diseases) (Suppl. Figures 2-6,10). By contrast, non-vaccine preventable respiratory infections including chickenpox (not part of routine vaccinations in the UK), scarlet fever, and streptococcal pharyngitis displayed an immediate resurgence after the removal of prevention measures (Suppl. Figures 1,7,8), suggesting that similar transmission peaks have been prevented by the vaccine-mediated immunity for the diseases with high vaccine coverage in the UK.

Concerns have also been raised about whether a lack of exposure to common pathogens may result in decreased immunity enabling larger and more deleterious outbreaks after the lifting of restrictions [3]. However, only four infectious diseases (chickenpox, herpes simplex virus, Skin and Subcutaneous Tissue Infections, Infectious Intestinal Diseases) have since the removal of all restrictions in England on 19^th^ July 2021 resulted in spread levels that exceeded those commonly observed pre-COVID-19 (Figure 1), and it remains to be investigated whether the observed increases may be related to COVID-19.

In conclusion, our analysis of the transmission patterns of infectious diseases shows that the COVID-19 prevention measures reduced, in particular, the spread of pathogens that are transmitted via the air and the faecal-oral route. Moreover, the COVID-19 prevention measures resulted in the sustained suppression of vaccine-preventable infectious diseases also after the removal of restrictions, while non-vaccine preventable diseases displayed a rapid rebound, supporting the importance of effective vaccination programmes. Despite concerns that a lack of exposure to common pathogens may affect population immunity and result in large outbreaks by various pathogens post-COVID-19, only four of the 22 investigated diseases and disease groups displayed higher post-than pre-pandemic levels without an obvious causative relationship. This included chickenpox for which an effective vaccine is available [15] but not used in the UK.

## Data Availability

All data produced in the present work are contained in the manuscript.

## Data availability statement

All data are provided in the manuscript and its supplements.

## References

1) Andrés P, Blandine P, Victoria D, William M, Justine O, Laurent E, Cedrine M, Bruno L, Aurelien T, Thomas J, Sophie TA, Manuel RC, Olivier T. Interactions between Severe Acute Respiratory Syndrome Coronavirus 2 Replication and Major Respiratory Viruses in Human nasal Epithelium. J Infect Dis. 2022 Aug 29:jiac357. doi: 10.1093/infdis/jiac357.

2) Eldesouki RE, Uhteg K, Mostafa HH. The circulation of Non-SARS-CoV-2 respiratory viruses and coinfections with SARS-CoV-2 during the surge of the Omicron variant. J Clin Virol. 2022 Aug;153:105215. doi: 10.1016/j.jcv.2022.105215.

3) Oh KB, Doherty TM, Vetter V, Bonanni P. Lifting non-pharmaceutical interventions following the COVID-19 pandemic - the quiet before the storm? Expert Rev Vaccines. 2022 Sep 5:1–13. doi: 10.1080/14760584.2022.2117693.

4) Shi HJ, Kim NY, Eom SA, Kim-Jeon MD, Oh SS, Moon BS, Kwon MJ, Eom JS. Effects of Non-Pharmacological Interventions on Respiratory Viruses Other Than SARS-CoV-2: Analysis of Laboratory Surveillance and Literature Review From 2018 to 2021. J Korean Med Sci. 2022 May 30;37(21):e172. doi: 10.3346/jkms.2022.37.e172.

5) Segala FV, Bavaro DF, Di Gennaro F, Salvati F, Marotta C, Saracino A, Murri R, Fantoni M. Impact of SARS-CoV-2 Epidemic on Antimicrobial Resistance: A Literature Review. Viruses. 2021 Oct 20;13(11):2110. doi: 10.3390/v13112110.

6) McCormick DW, Kugeler KJ, Marx GE, Jayanthi P, Dietz S, Mead P, Hinckley AF. Effects of COVID-19 Pandemic on Reported Lyme Disease, United States, 2020. Emerg Infect Dis. 2021 Oct;27(10):2715–2717. doi: 10.3201/eid2710.210903.

7) Piotrowski M, Rymaszewska A. The Impact of a Pandemic COVID-19 on the Incidence of Borreliosis in Poland. Acta Parasitol. 2022 Jun;67(2):1007–1009. doi: 10.1007/s11686-021-00495-0.

8) Kim S, Kim J, Choi BY, Park B. Trends in gastrointestinal infections before and during non-pharmaceutical interventions in Korea in comparison with the United States. Epidemiol Health. 2022;44:e2022011. doi: 10.4178/epih.e2022011.

9) Nielsen RT, Dalby T, Emborg HD, Larsen AR, Petersen A, Torpdahl M, Hoffmann S, Vestergaard LS, Valentiner-Branth P. COVID-19 preventive measures coincided with a marked decline in other infectious diseases in Denmark, spring 2020. Epidemiol Infect. 2022 Jul 28;150:e138. doi: 10.1017/S0950268822001145.

10) Boom WH, Schaible UE, Achkar JM. The knowns and unknowns of latent Mycobacterium tuberculosis infection. J Clin Invest. 2021 Feb 1;131(3):e136222. doi: 10.1172/JCI136222.

11) Trajman A, Felker I, Alves LC, Coutinho I, Osman M, Meehan SA, Singh UB, Schwartz Y. The COVID-19 and TB syndemic: the way forward. Int J Tuberc Lung Dis. 2022 Aug 1;26(8):710–719. doi: 10.5588/ijtld.22.0006.

12) Wang Q, Guo S, Wei X, Dong Q, Xu N, Li H, Zhao J, Sun Q. Global prevalence, treatment and outcome of tuberculosis and COVID-19 coinfection: a systematic review and meta-analysis (from November 2019 to March 2021). BMJ Open. 2022 Jun 20;12(6):e059396. doi: 10.1136/bmjopen-2021-059396.

13) Knox MA, Garcia-R JC, Ogbuigwe P, Pita A, Velathanthiri N, Hayman DTS. Absence of Cryptosporidium hominis and dominance of zoonotic Cryptosporidium species in patients after Covid-19 restrictions in Auckland, New Zealand. Parasitology. 2021 Sep;148(11):1288–1292. doi: 10.1017/S0031182021000974.

14) Lucero Y, Matson DO, Ashkenazi S, George S, O’Ryan M. Norovirus: Facts and Reflections from Past, Present, and Future. Viruses. 2021 Nov 30;13(12):2399. doi: 10.3390/v13122399.

15) Otani N, Shima M, Yamamoto T, Okuno T. Effect of Routine Varicella Immunization on the Epidemiology and Immunogenicity of Varicella and Shingles. Viruses. 2022 Mar 12;14(3):588. doi: 10.3390/v14030588.

