## Supplemental information for "Impact of the COVID-19 pandemic on the circulation of other pathogens in England"

**Running title:**

Lauren Hayes[1#], Hannah Uri[1#], Denisa Bojkova[2], Jindrich Cinatl jr.[2,3], Mark N. Wass[1]*, Martin Michaelis[1]*

[1] School of Biosciences, University of Kent, Canterbury CT2 7NJ, UK

[2] Institute for Medical Virology, University Hospital, Goethe University, Paul Ehrlich-Str. 40, 60596 Frankfurt am Main, Germany

[3] Dr. Petra Joh-Forschungshaus, Komturstr. 3A, 60528 Frankfurt am Main, Germany

[#] equal contribution

* Corresponding authors:

Mark Wass, School of Biosciences, University of Kent, Canterbury CT2 7NJ, UK; phone +44 1227 82 7626;

Martin Michaelis, School of Biosciences, University of Kent, Canterbury CT2 7NJ, UK; phone +44 1227 82 7804;

**Suppl. Methods**

**Infectious disease data sources**

The infectious diseases that were studied in this report are presented together with their anticipated modes of transmission in Suppl. Table 1. ‘Influenza-like illness’ include diseases that were diagnosed as influenza based on the sudden onset of clinical symptoms including runny nose, fever, malaise, aches, cough, sneezing, and nausea, which was not always confirmed by a diagnostic test. Hence, some cases may have been caused by other respiratory viruses [Fitzner et al., 2018]. ‘Skin and subcutaneous tissue infections’ encompasses both uncomplicated and necrotising pathological conditions of the skin or subcutaneous fat, resulting in erythema, oedema, inflammation, and pain. This includes (but is not limited to) diseases such as cellulitis, impetigo, folliculitis, abscesses, carbuncles, and trauma-related infections [Esposito et al., 2017]. ‘Infectious intestinal diseases’ refers to infections of the stomach, small intestine, and/or bowel, with symptoms including diarrhoea, vomiting, and abdominal pain, typically reflecting diseases such as gastroenteritis, cholera, and typhoid fever [Donaldson et al., 2019].

Weekly case numbers were available for England for all diseases, except for methicillin resistant *Staphylococcus aureus* (MRSA), Lyme disease, and hepatitis E that were recorded quarterly. For diseases with seasonal transmission patterns (influenza-like illnesses, pneumococcal disease, strep throat, scarlet fever cryptosporidiosis, foodborne illness, norovirus, Lyme disease), the average season peaks were calculated based on the included pre-COVID-19 years.

Infectious disease case number dynamics during the COVID-19 pandemic were compared to those of COVID-19 and in the context of the prevention measures that were in place at the time. Rubella was excluded from the analysis due to low case numbers (<5) and diseases with cumulative quarterly cases due to lack of comparable data.

Measles, mumps and rubella (MMR), tuberculosis, scarlet fever, foodborne illness and whooping cough case numbers reported to the UK Health Security Agency (UKHSA) by medical practitioners were derived from the PHE Notifications of Infectious Diseases (NOIDs) database [https://www.gov.uk/government/collections/notifications-of-infectious-diseases-noids]. Lyme disease, hepatitis C, hepatitis E, cryptosporidiosis, shigellosis, strep throat, and pneumococcal disease case numbers were derived from NOIDs causative agent reports, which are based on notifications from laboratories in England [https://www.gov.uk/government/collections/notifications-of-infectious-diseases-noids].

MRSA data was derived from the joint UKHSA/ Office for National Statistics (ONS) MRSA bacteraemia monthly count reports [https://www.gov.uk/government/statistics/mrsa-bacteraemia-monthly-data-by-location-of-onset].

Laboratory confirmed norovirus cases were extracted from 2020 to 2022 national norovirus and rotavirus bulletins (from 2020 onwards) [https://www.gov.uk/government/statistical-data-sets/national-norovirus-and-rotavirus-bulletin-management-information--2] and weekly UKHSA reports (prior to 2020) [https://www.gov.uk/government/statistics/norovirus-and-rotavirus-summary-of-surveillance-2019-to-2020]. Data was extracted from the norovirus routine laboratory reports of positive norovirus samples from the Second Generation Surveillance System (SGSS).

Chickenpox, influenza-like-illness (ILI), herpes simplex virus (HSV), infectious intestinal disease, skin and subcutaneous tissue infection (SSTI), and urinary tract infection (UTI) data was derived from the Royal Collage of General Practitioners (RCGP) Research and Surveillance Centre (RSC) public health data [https://www.rcgp.org.uk/representing-you/research-at-rcgp/research-surveillance-centre/public-health-data]. COVID-19 data was derived from the Coronavirus (COVID-19) Infection Survey of the ONS [https://www.ons.gov.uk/peoplepopulationandcommunity/healthandsocialcare/conditionsanddiseases/datasets/coronaviruscovid19infectionsurveydata]. Information on circulating SARS-CoV-2 variants was derived from the variants of concern technical briefing 44 (22 July 2022) of UKHSA [https://www.gov.uk/government/publications/investigation-of-sars-cov-2-variants-technical-briefings].

**Timeline of protection measures**

An overview of the timing of prevention measures is provided in Suppl. Table 2. Dates and guidance are based on the British Foreign Policy Group (BFPG) COVID-19 Timeline by Evie Aspinall (https://bfpg.co.uk/2020/04/covid-19-timeline/). The accuracy of this information was confirmed using GOV.UK guidance, policy papers, and records including prime minister statements and daily press briefings on governmental responses to the COVID-19 pandemic (Prime Minister's statement on coronavirus (COVID-19): 23 March 2020 [Internet]. GOV.UK. [cited 2022 Aug 16]. Available from: https://www.gov.uk/government/speeches/pm-address-to-the-nation-on-coronavirus-23-march-2020; https://www.gov.uk/government/speeches/pm-statement-on-coronavirus-18-march-2020; https://www.gov.uk/government/news/prime-minister-announces-new-national-restrictions; https://lordslibrary.parliament.uk/covid-19-local-alert-levels-three-tier-system-for-england; https://www.gov.uk/government/publications/covid-19-response-autumn-and-winter-plan-2021; https://www.gov.uk/government/publications/covid-19-response-spring-2021; https://www.gov.uk/government/news/prime-minister-confirms-move-to-step-4).

### References

Donaldson AL, Clough HE, O'Brien SJ, Harris JP. Symptom profiling for infectious intestinal disease (IID): a secondary data analysis of the IID2 study. Epidemiol Infect. 2019 Jan;147:e229. doi: 10.1017/S0950268819001201.

Esposito S, Bassetti M, Concia E, De Simone G, De Rosa FG, Grossi P, Novelli A, Menichetti F, Petrosillo N, Tinelli M, Tumbarello M, Sanguinetti M, Viale P, Venditti M, Viscoli C; Italian Society of Infectious and Tropical Diseases. Diagnosis and management of skin and soft-tissue infections (SSTI). A literature review and consensus statement: an update. J Chemother. 2017 Aug;29(4):197-214. doi: 10.1080/1120009X.2017.1311398.

Fitzner J, Qasmieh S, Mounts AW, Alexander B, Besselaar T, Briand S, Brown C, Clark S, Dueger E, Gross D, Hauge S, Hirve S, Jorgensen P, Katz MA, Mafi A, Malik M, McCarron M, Meerhoff T, Mori Y, Mott J, Olivera MTDC, Ortiz JR, Palekar R, Rebelo-de-Andrade H, Soetens L, Yahaya AA, Zhang W, Vandemaele K. Revision of clinical case definitions: influenza-like illness and severe acute respiratory infection. Bull World Health Organ. 2018 Feb 1;96(2):122-128. doi: 10.2471/BLT.17.194514.

**Suppl. Figure 1**

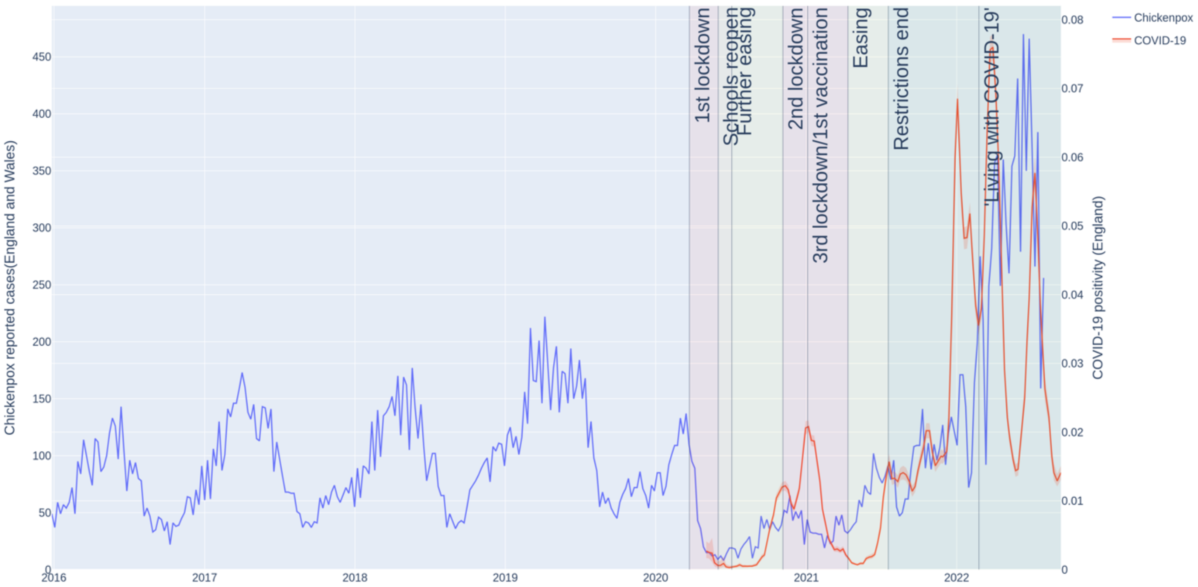


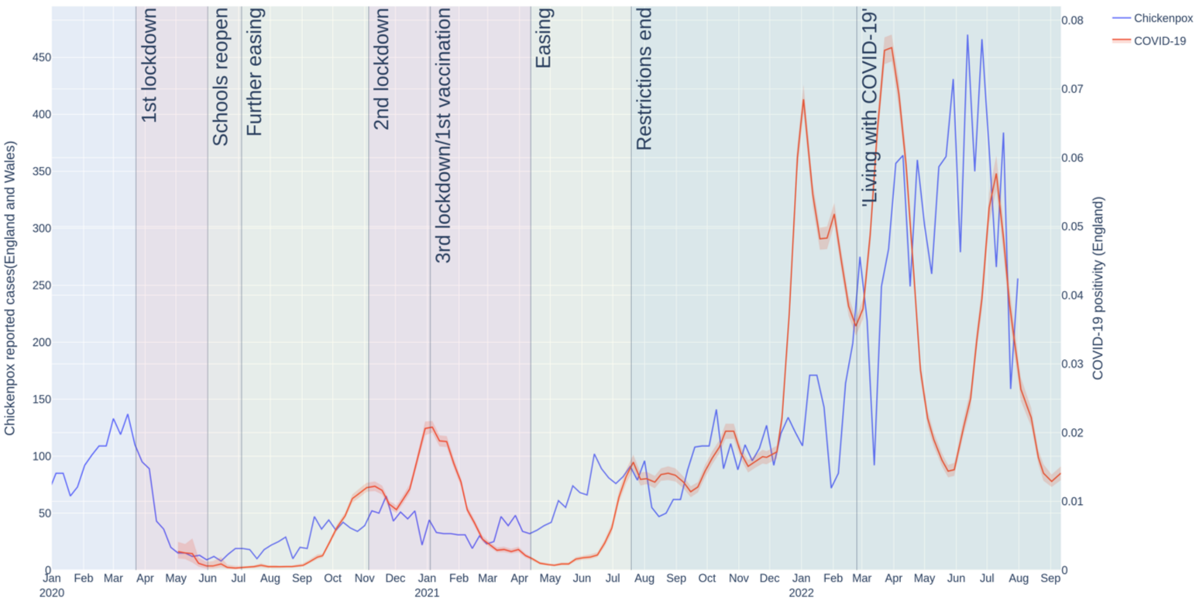


**Suppl. Figure 1. Chickenpox case numbers during the COVID-19 pandemic in England.** Weekly case numbers (chickenpox: left y-axis, blue line, COVID-19: right y-axis, orange line) starting from 1st January 2017 (top graph) or 30th December 2019 (bottom graph).

**Suppl. Figure 2**

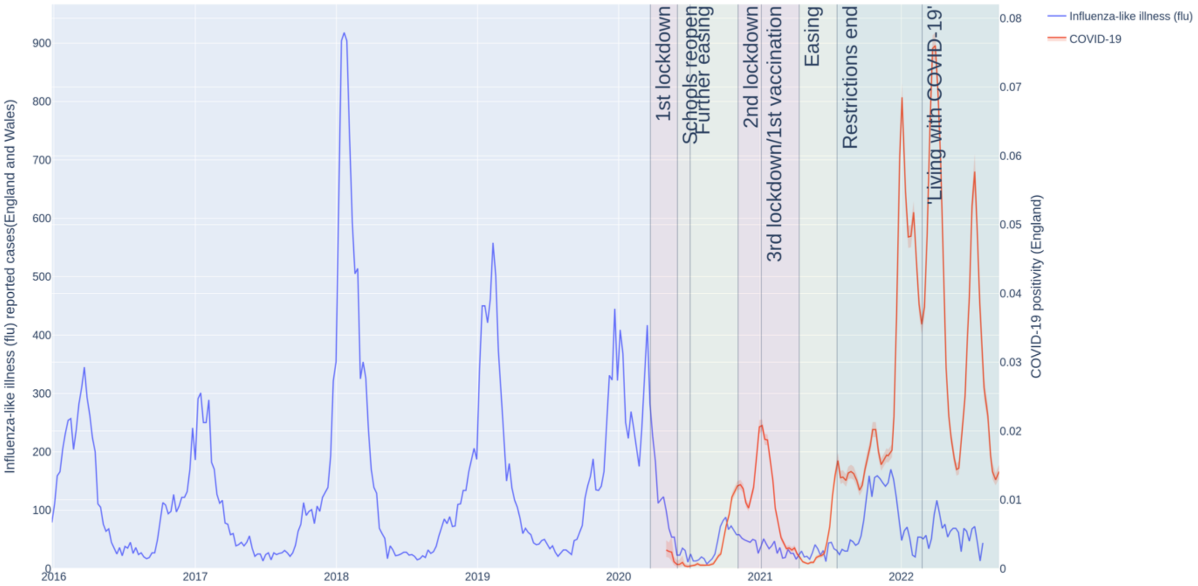


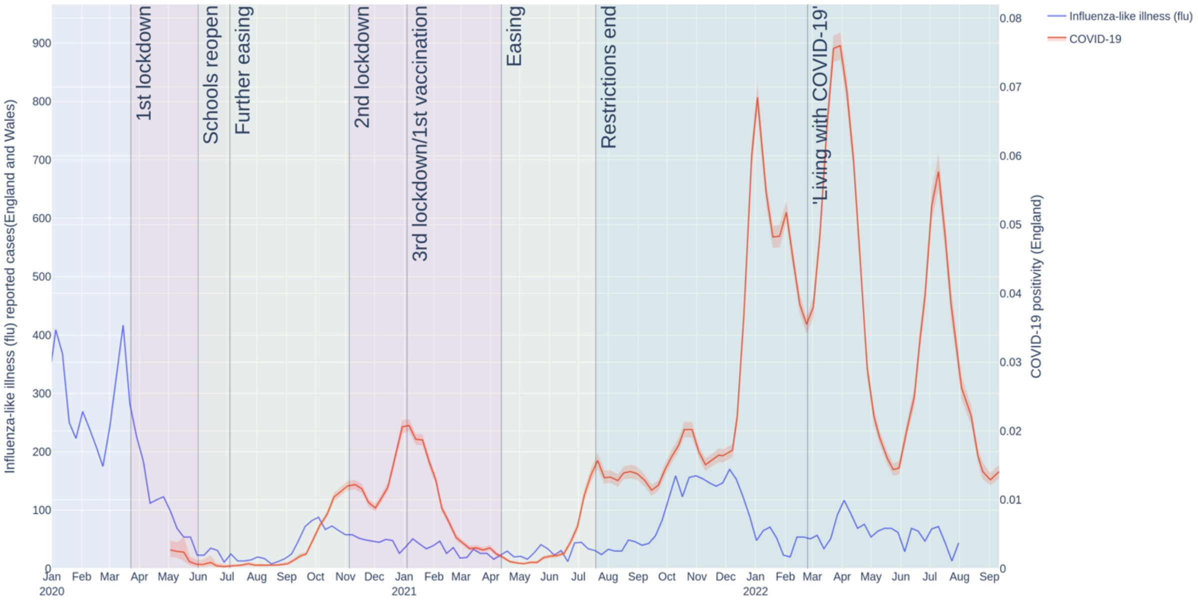


**Suppl. Figure 2. Influenza-like illness case numbers during the COVID-19 pandemic in England.** Weekly case numbers (influenza-like illnesses: left y-axis, blue line, COVID-19: right y-axis, orange line) starting from 1st January 2016 (top graph) or 30th December 2019 (bottom graph).

**Suppl. Figure 3**

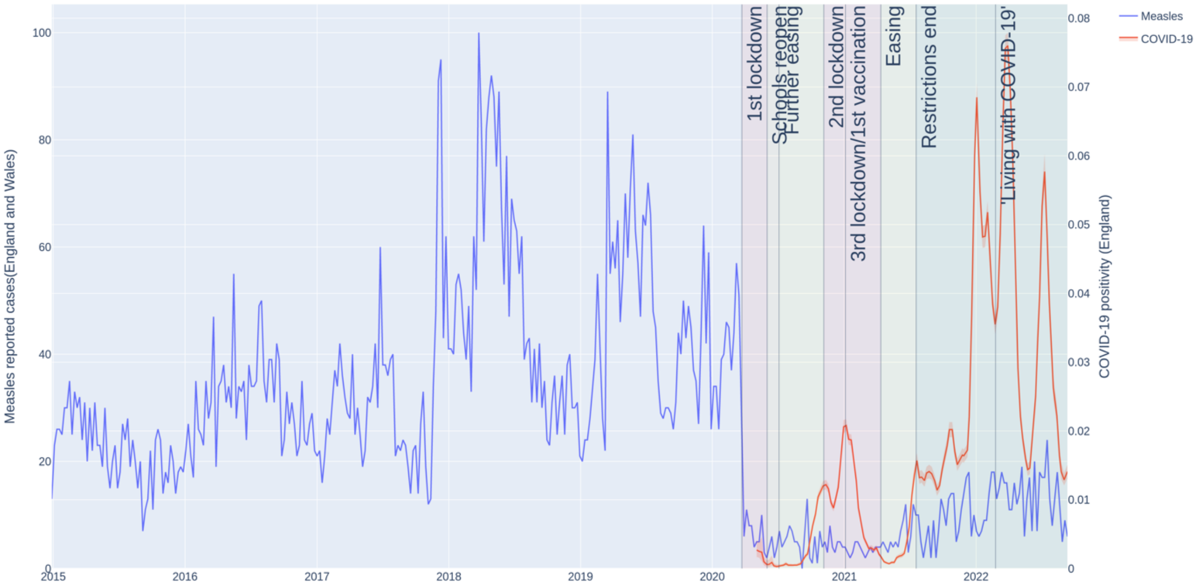


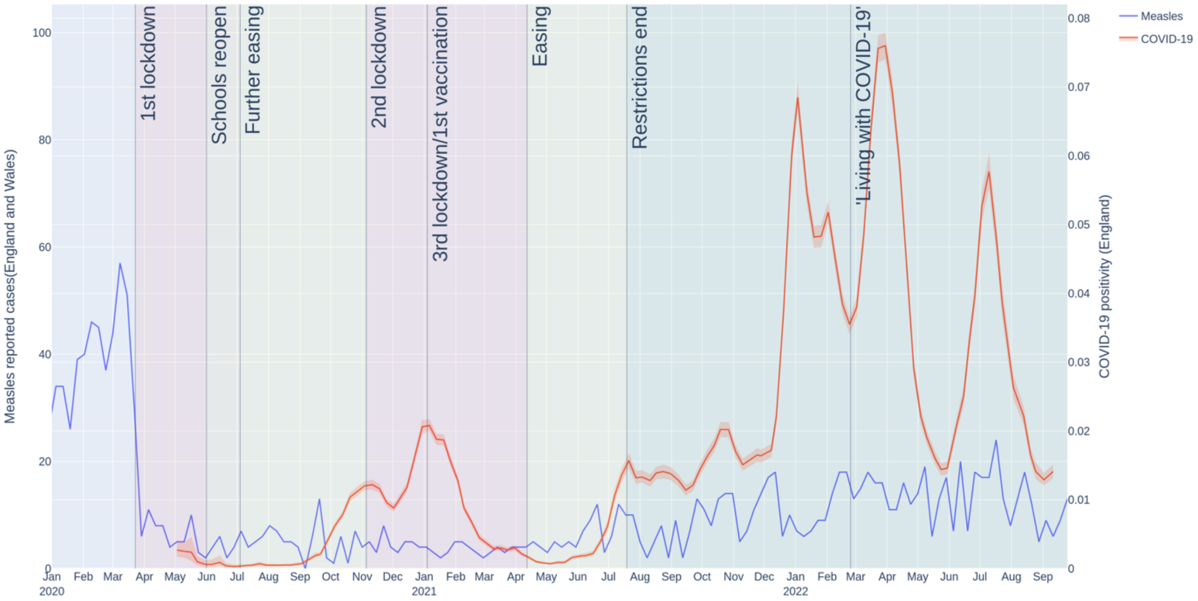


**Suppl. Figure 3. Measles case numbers during the COVID-19 pandemic in England.** Weekly case numbers (measles: left y-axis, blue line, COVID-19: right y-axis, orange line) starting from 1st January 2015 (top graph) or 30th December 2019 (bottom graph).

**Suppl. Figure 4**

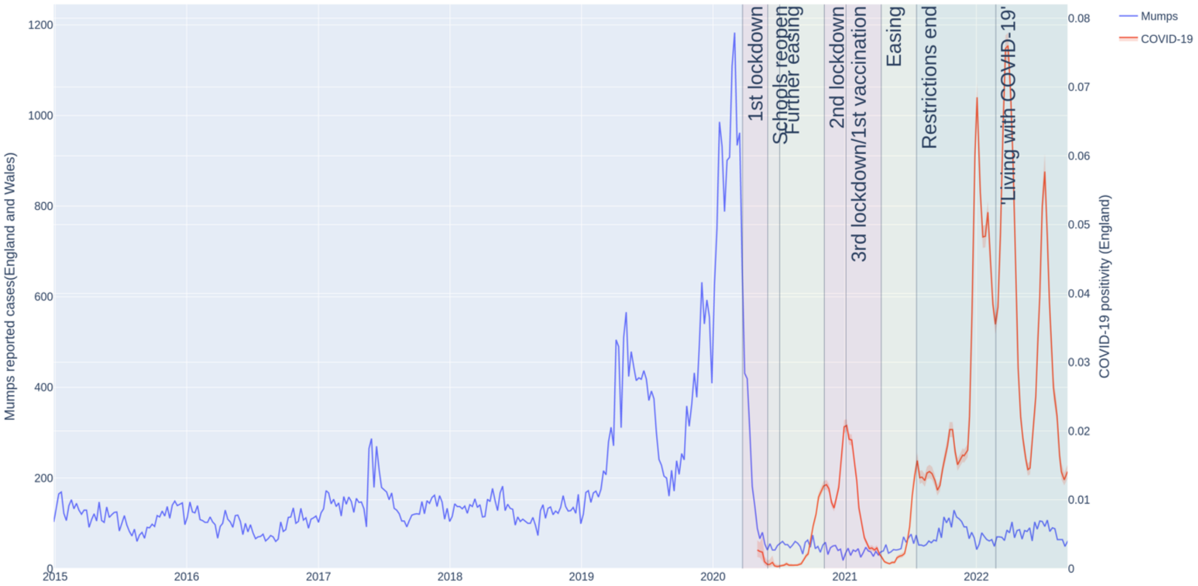


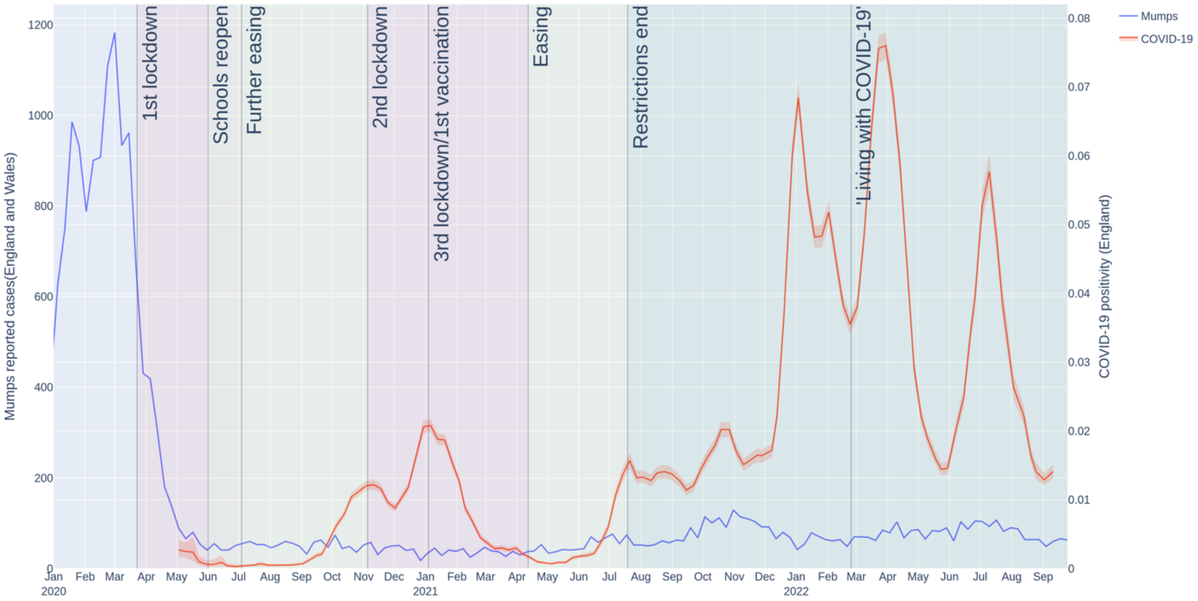


**Suppl. Figure 4. Mumps case numbers during the COVID-19 pandemic in England.** Weekly case numbers (mumps: left y-axis, blue line, COVID-19: right y-axis, orange line) starting from 1st January 2015 (top graph) or 30th December 2019 (bottom graph).

**Suppl. Figure 5**

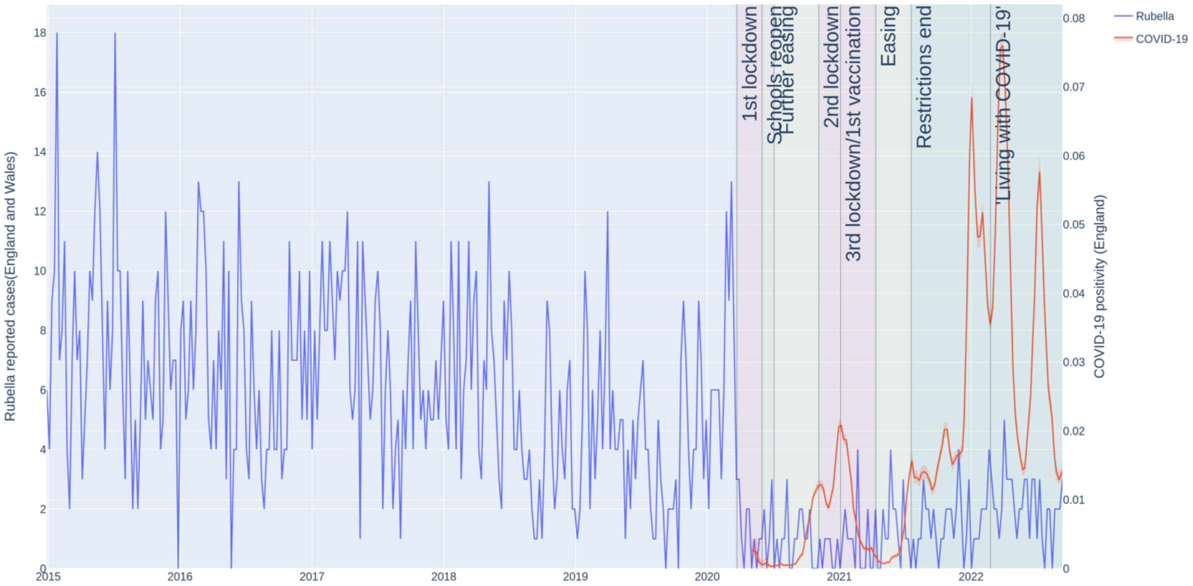


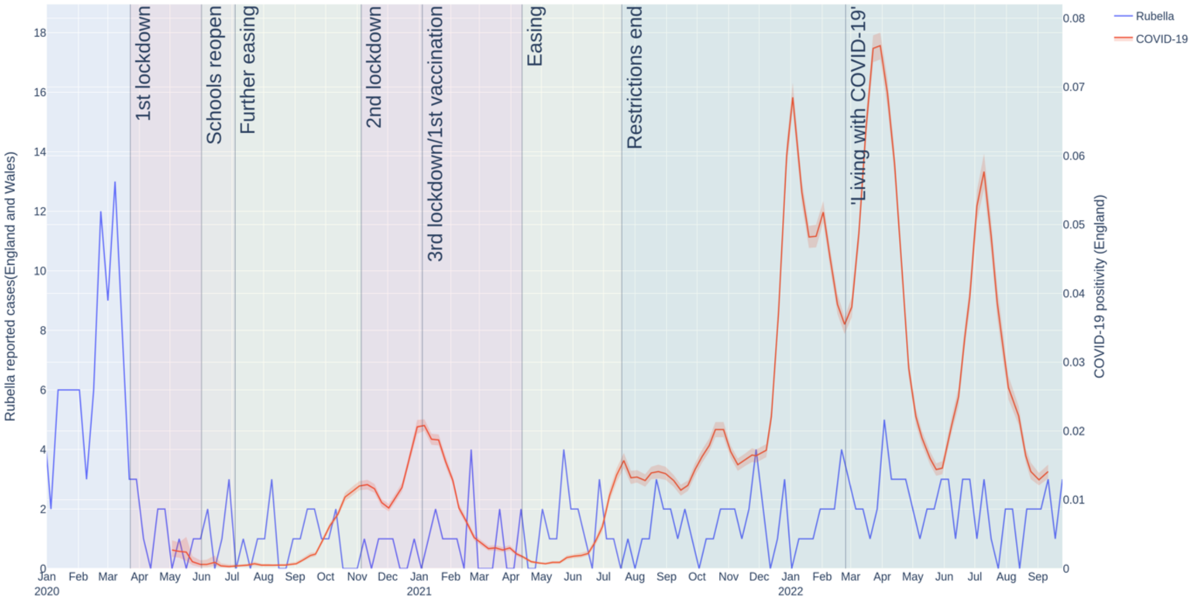


**Suppl. Figure 5. Rubella case numbers during the COVID-19 pandemic in England.** Weekly case numbers (rubella: left y-axis, blue line, COVID-19: right y-axis, orange line) starting from 1st January 2015 (top graph) or 30th December 2019 (bottom graph).

**Suppl. Figure 6**

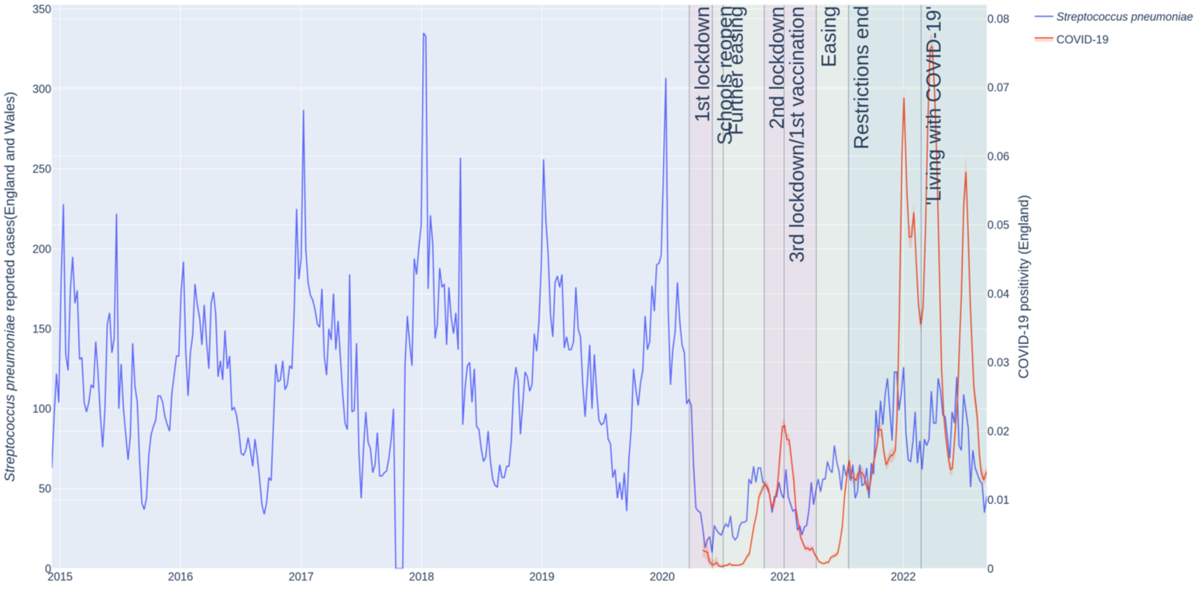


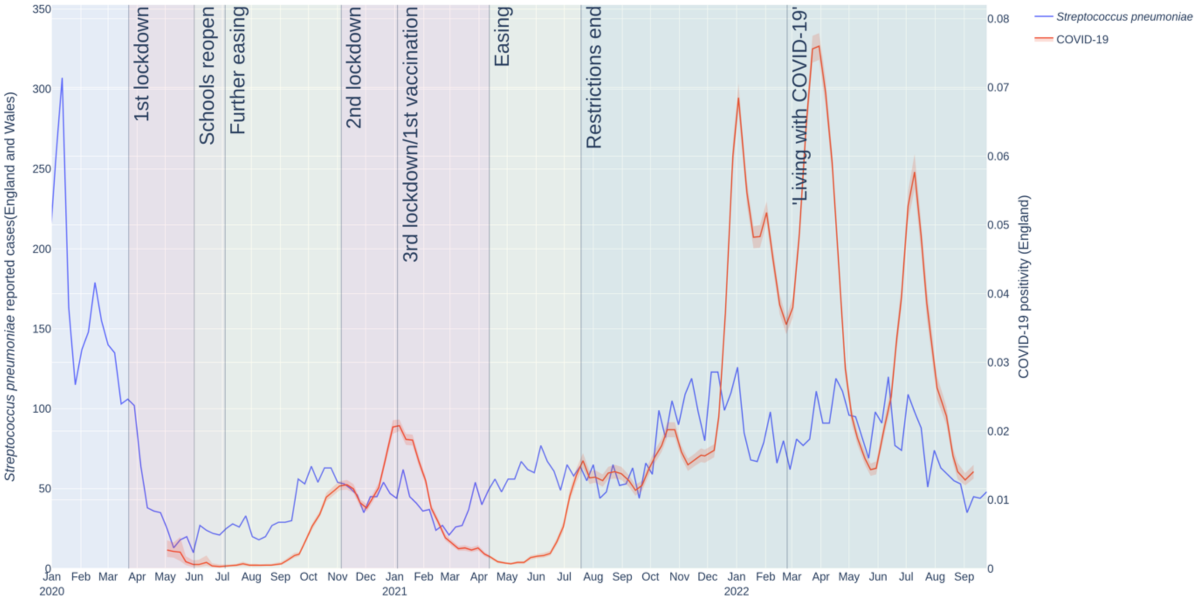


**Suppl. Figure 6. Pneumococcal disease (*Streptococcus pneumoniae*) case numbers during the COVID-19 pandemic in England.** Weekly case numbers (*Streptococcus pneumoniae*: left y-axis, blue line, COVID-19: right y-axis, orange line) starting from 1st January 2015 (top graph) or 30th December 2019 (bottom graph).

**Suppl. Figure 7**

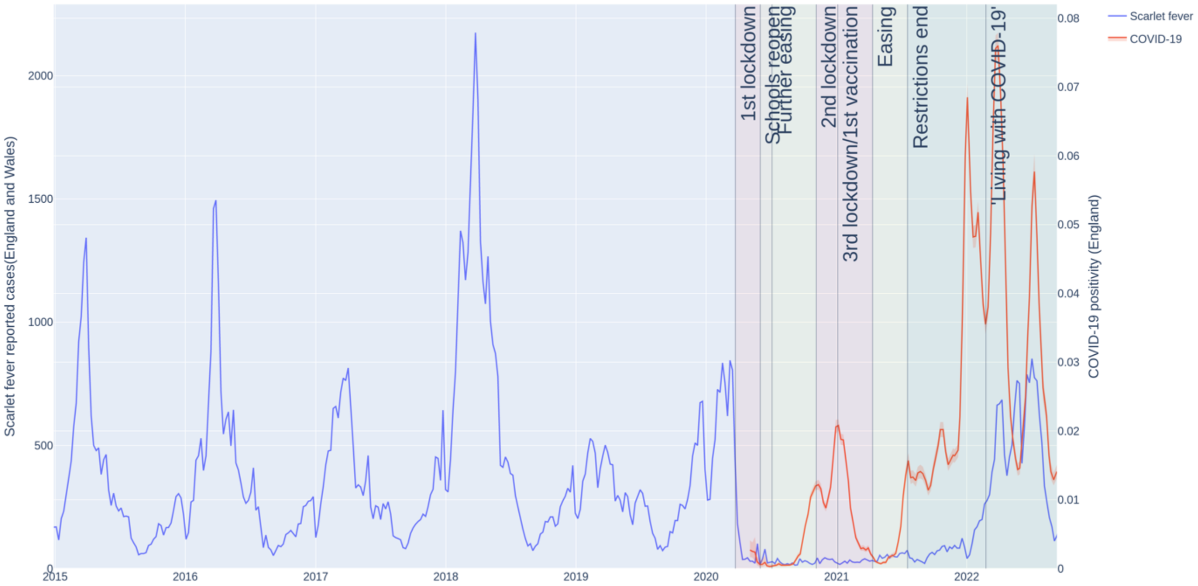


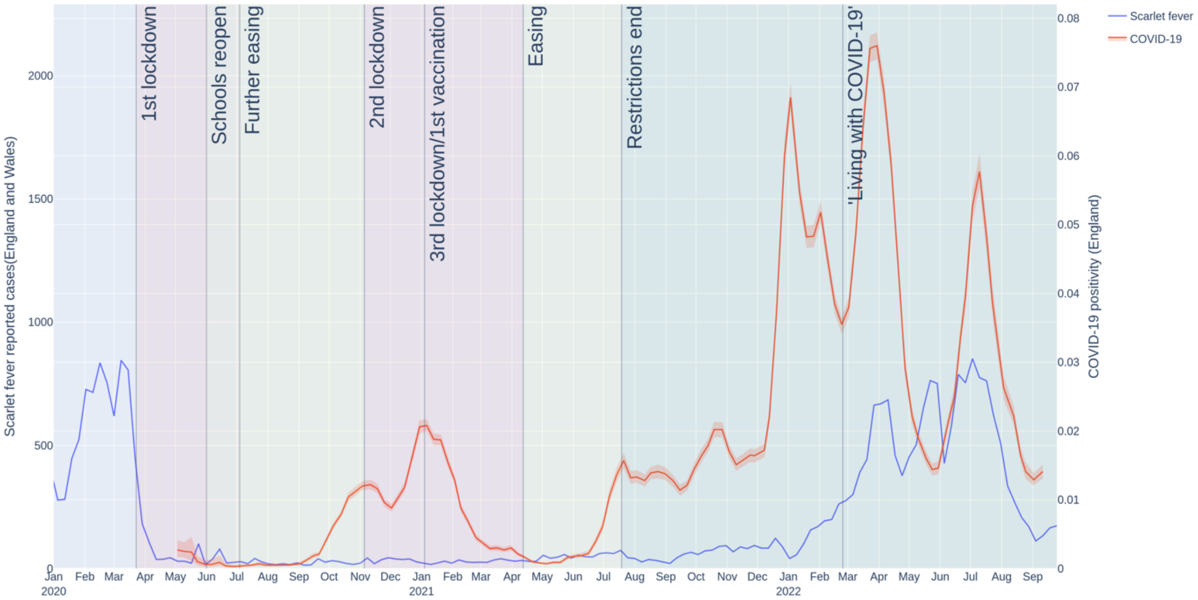


**Suppl. Figure 7. Scarlet fever case numbers during the COVID-19 pandemic in England.** Weekly case numbers (scarlet fever: left y-axis, blue line, COVID-19: right y-axis, orange line) starting from 1st January 2015 (top graph) or 30th December 2019 (bottom graph).

**Suppl. Figure 8**

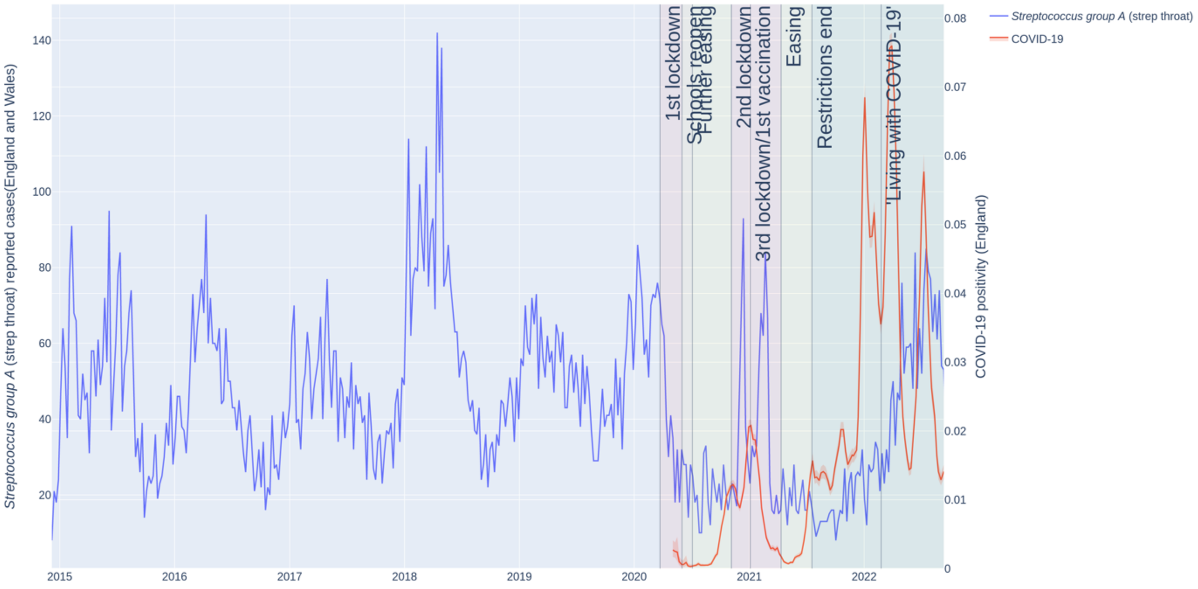


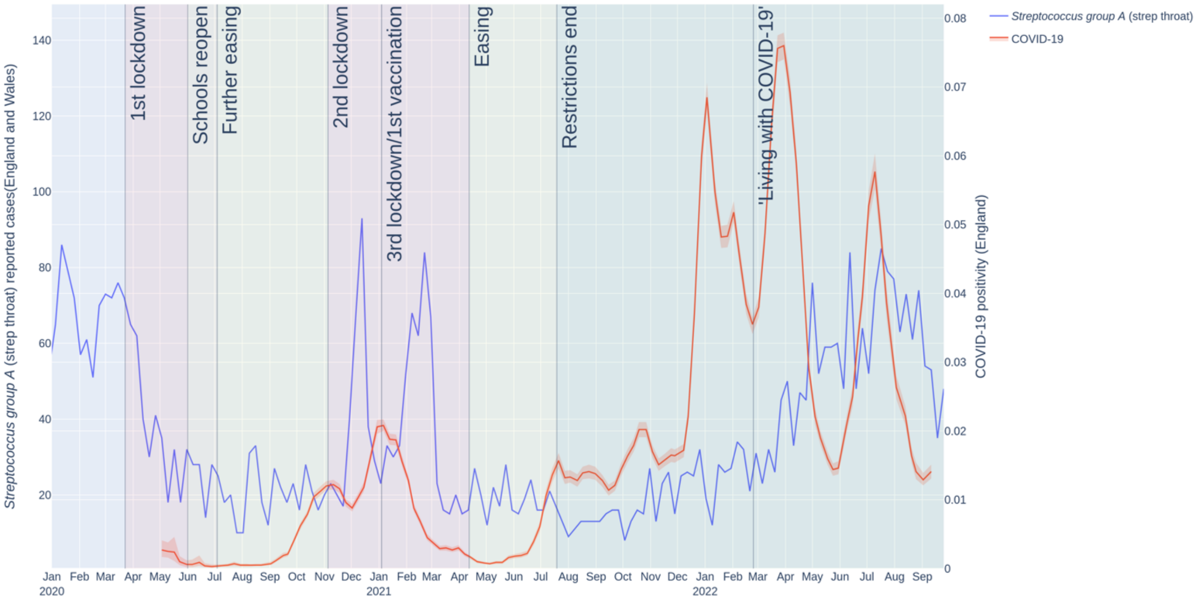


**Suppl. Figure 8. Streptococcal pharyngitis (Streptococcus group A, strep throat) case numbers during the COVID-19 pandemic in England.** Weekly case numbers (Streptococcus group A: left y-axis, blue line, COVID-19: right y-axis, orange line) starting from 1st January 2015 (top graph) or 30th December 2019 (bottom graph).

**Suppl. Figure 9**

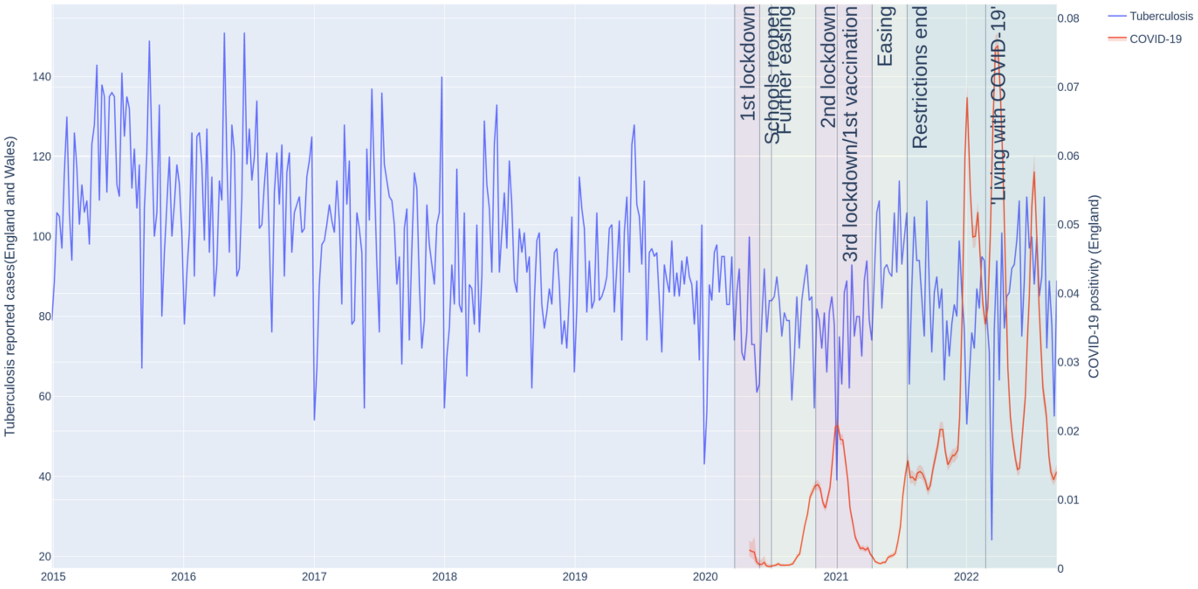


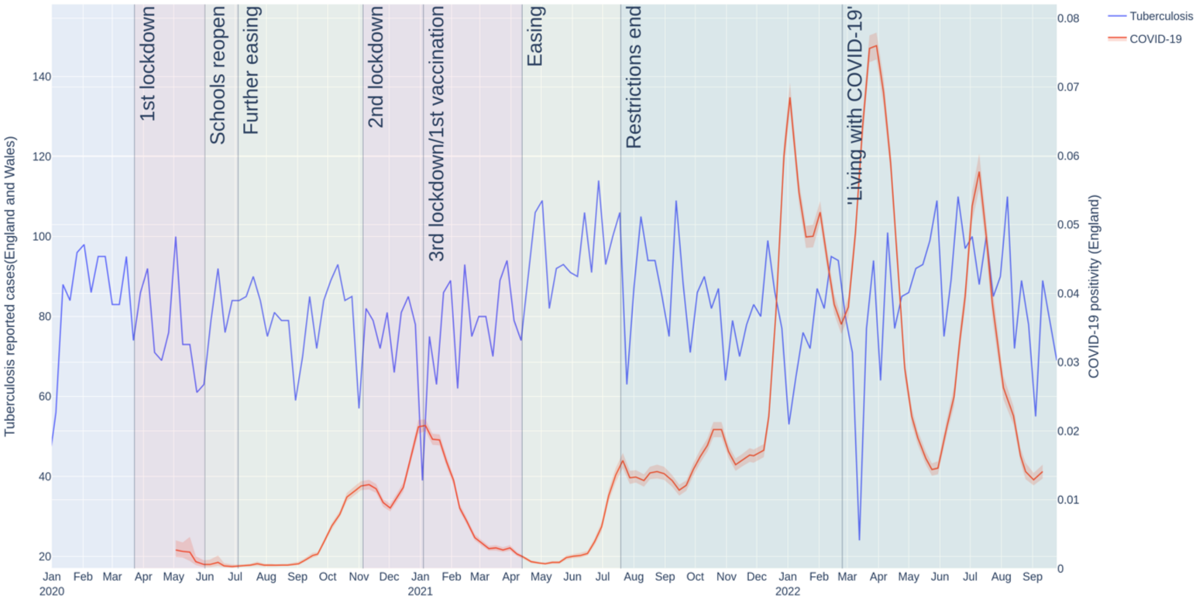


**Suppl. Figure 9. Tuberculosis case numbers during the COVID-19 pandemic in England.** Weekly case numbers (tuberculosis: left y-axis, blue line, COVID-19: right y-axis, orange line) starting from 1st January 2015 (top graph) or 30th December 2019 (bottom graph).

**Suppl. Figure 10**

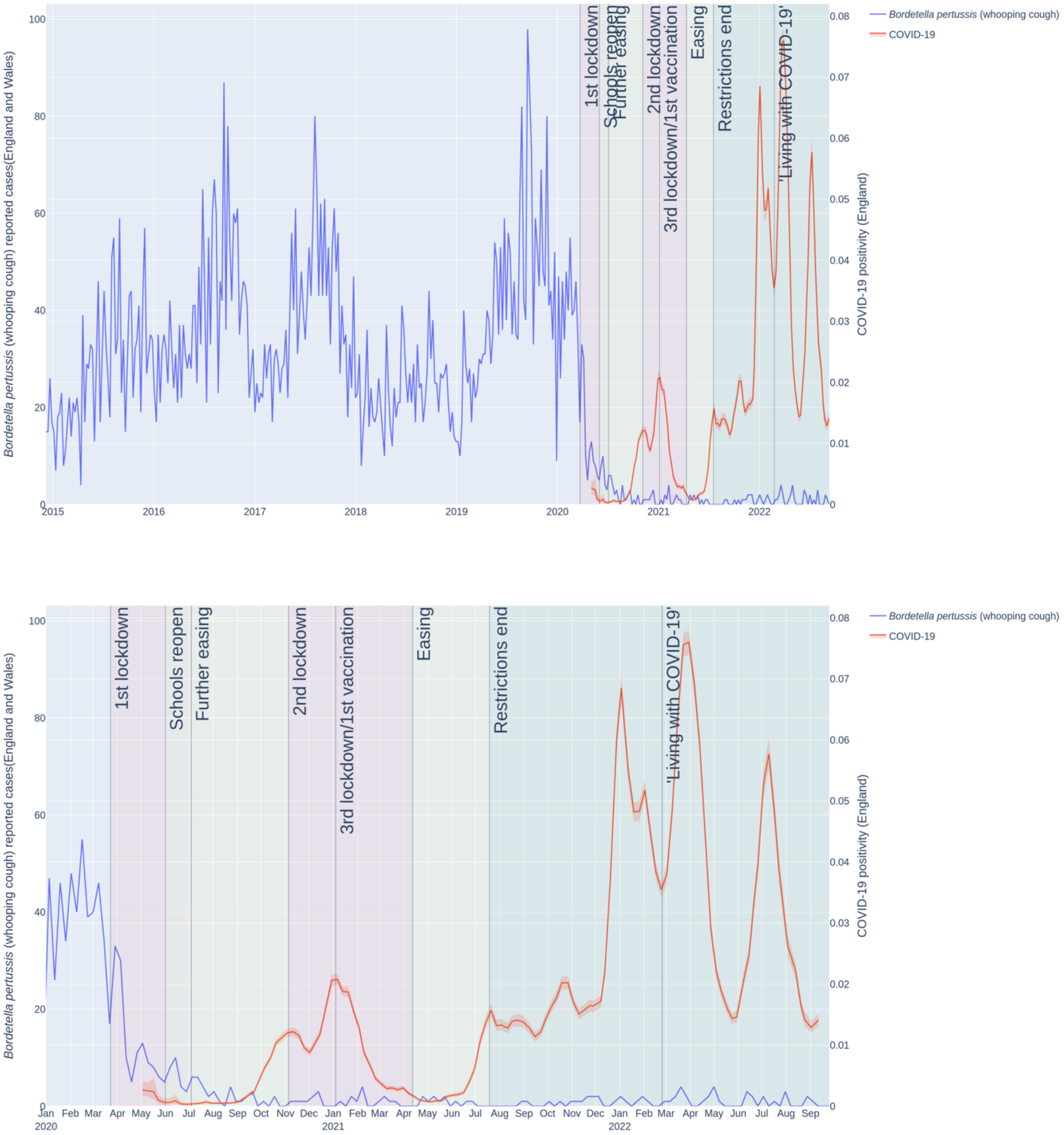


**Suppl. Figure 10. Pertussis (*Bordetella pertussis*, whooping cough) case numbers during the COVID-19 pandemic in England.** Weekly case numbers (pertussis: left y-axis, blue line, COVID-19: right y-axis, orange line) starting from 1st January 2015 (top graph) or 30th December 2019 (bottom graph).

**Suppl. Figure 11**

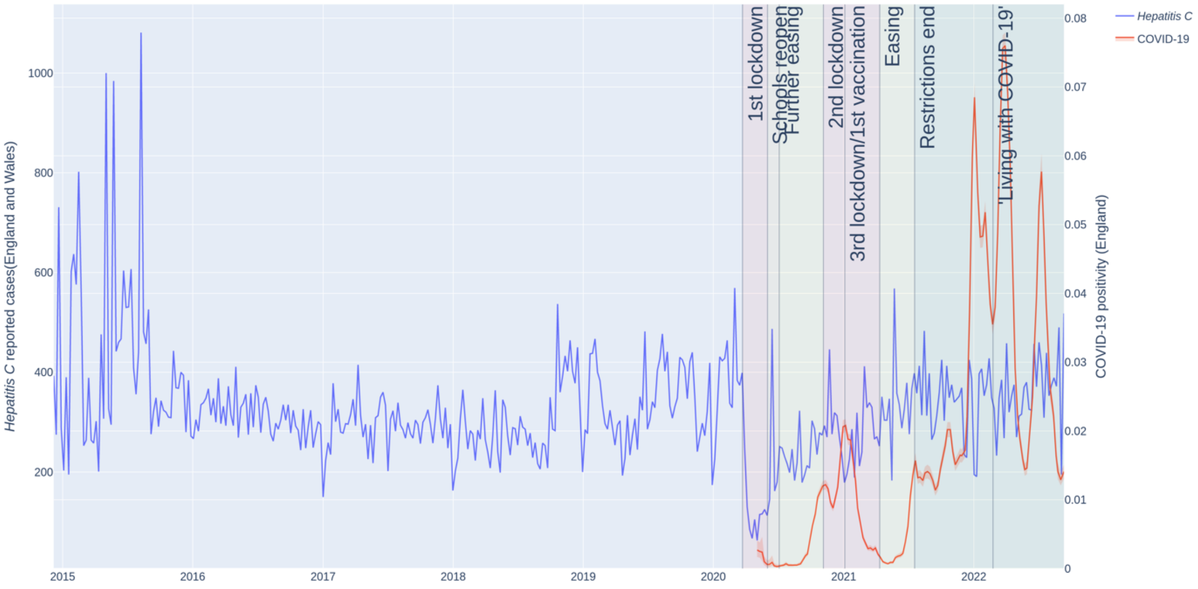


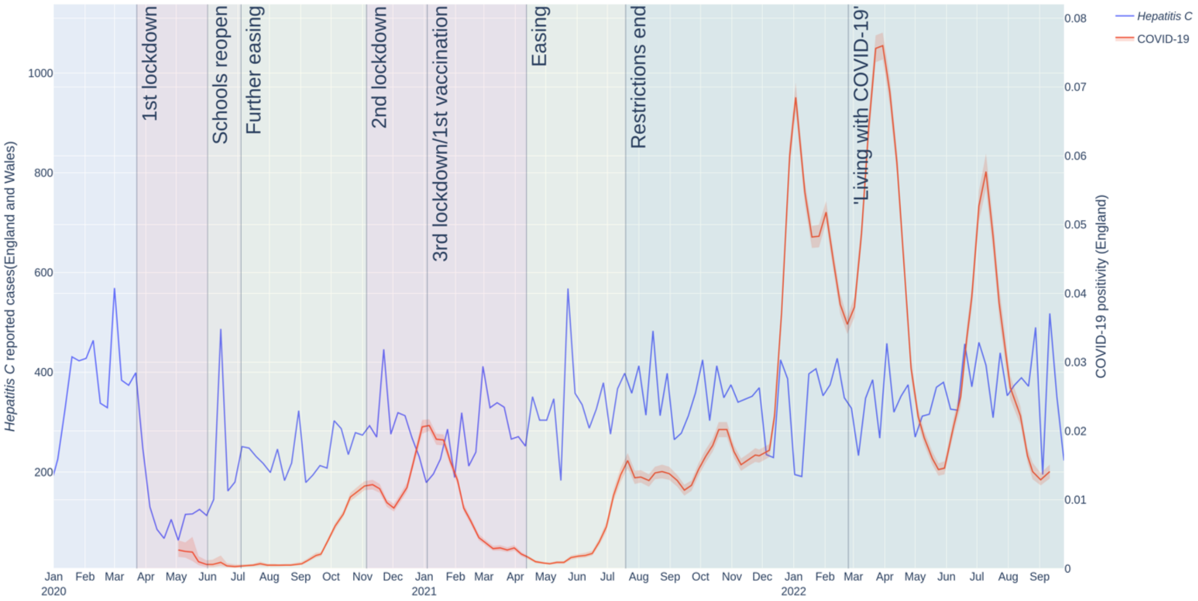


**Suppl. Figure 11. Hepatitis C case numbers during the COVID-19 pandemic in England.** Weekly case numbers (hepatitis C: left y-axis, blue line, COVID-19: right y-axis, orange line) starting from 1st January 2015 (top graph) or 30th December 2019 (bottom graph).

**Suppl. Figure 12**

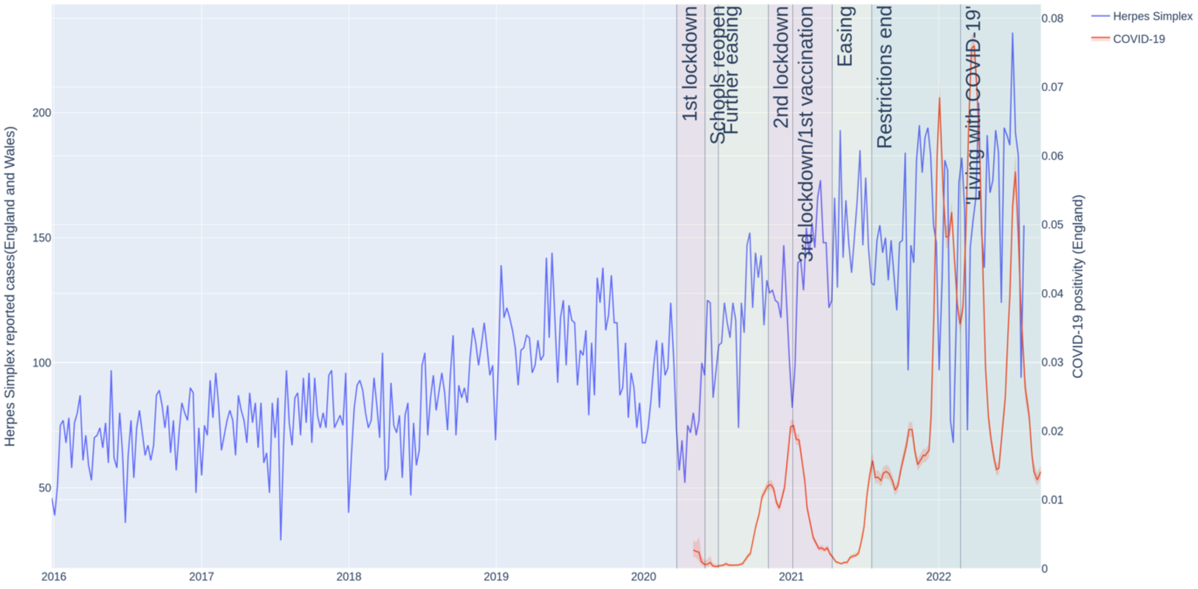


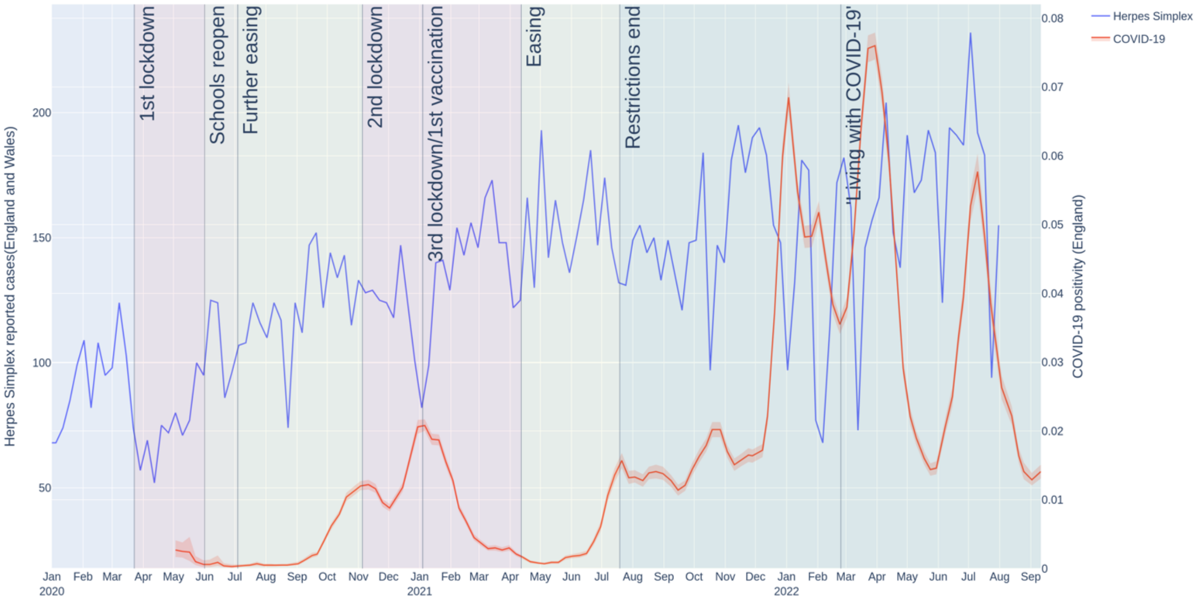


**Suppl. Figure 12. Herpes simplex virus case numbers during the COVID-19 pandemic in England.** Weekly case numbers (herpes simplex virus: left y-axis, blue line, COVID-19: right y-axis, orange line) starting from 1st January 2016 (top graph) or 30th December 2019 (bottom graph).

**Suppl. Figure 13**

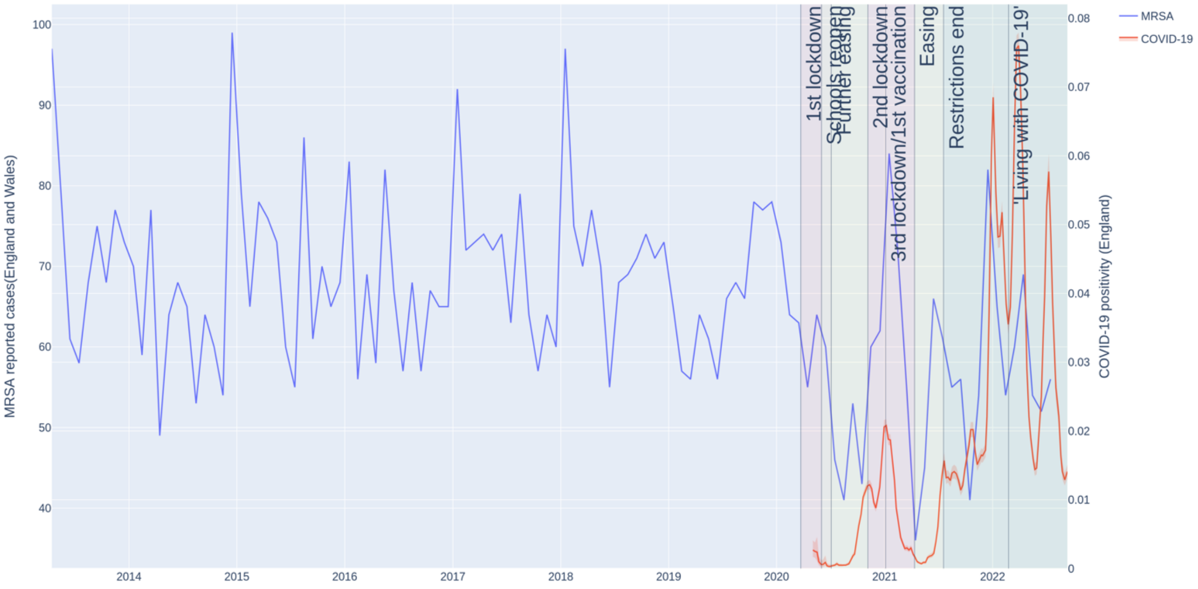


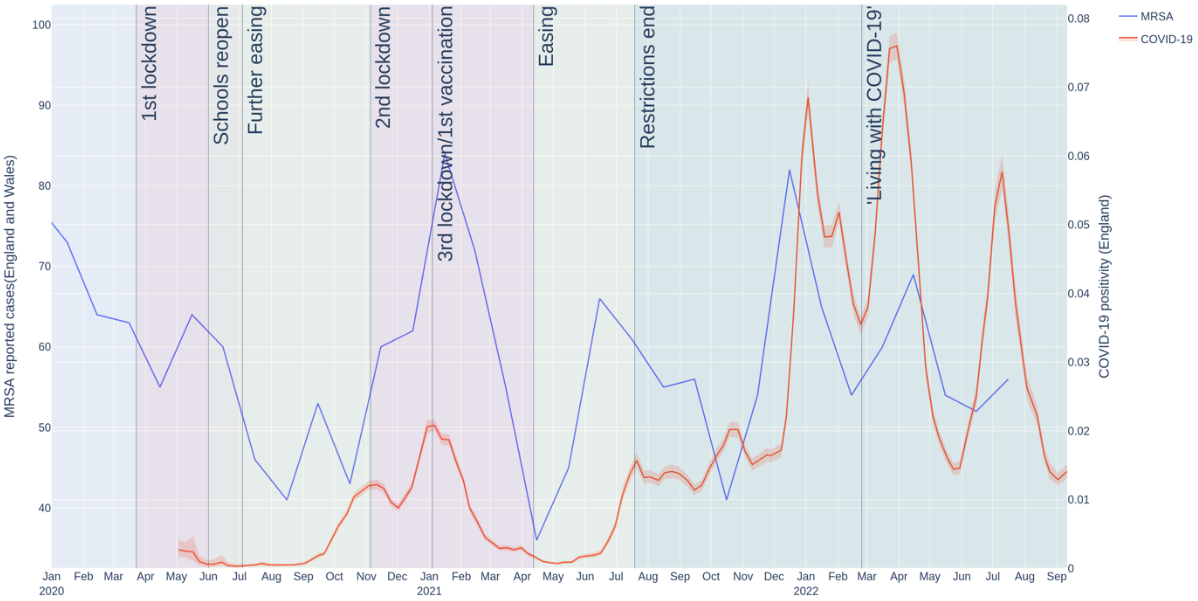


**Suppl. Figure 13. Methicillin-resistant *Staphylococcus aureus* (MRSA) case numbers during the COVID-19 pandemic in England.** Weekly case numbers (MRSA: left y-axis, blue line, COVID-19: right y-axis, orange line) starting from 1st January 2017 (top graph) or 30th December 2019 (bottom graph).

### Suppl. Figure 14

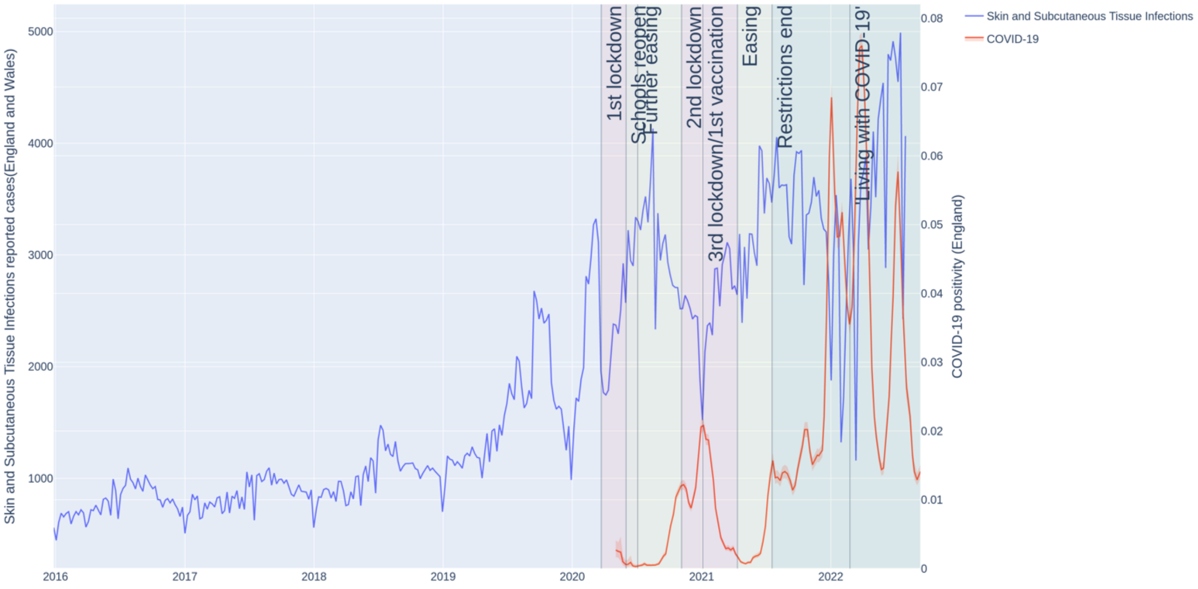


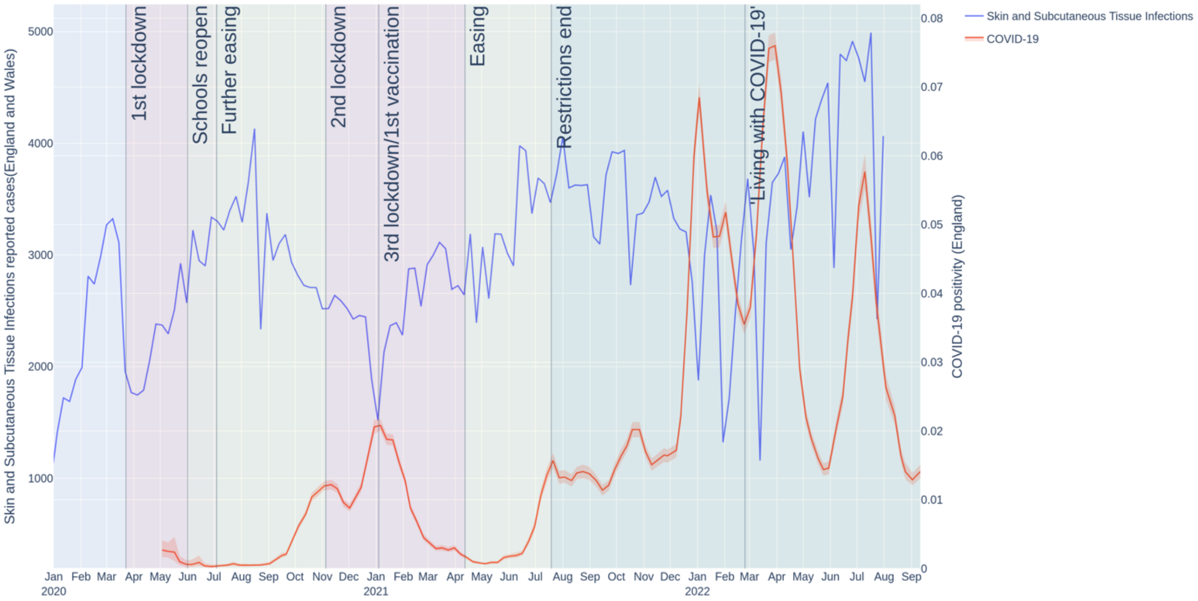


**Suppl. Figure 14. Skin and Subcutaneous Tissue Infections case numbers during the COVID-19 pandemic in England.** Weekly case numbers (Skin and Subcutaneous Tissue Infections: left y-axis, blue line, COVID-19: right y-axis, orange line) starting from 1st January 2016 (top graph) or 30th December 2019 (bottom graph).

**Suppl. Figure 15**

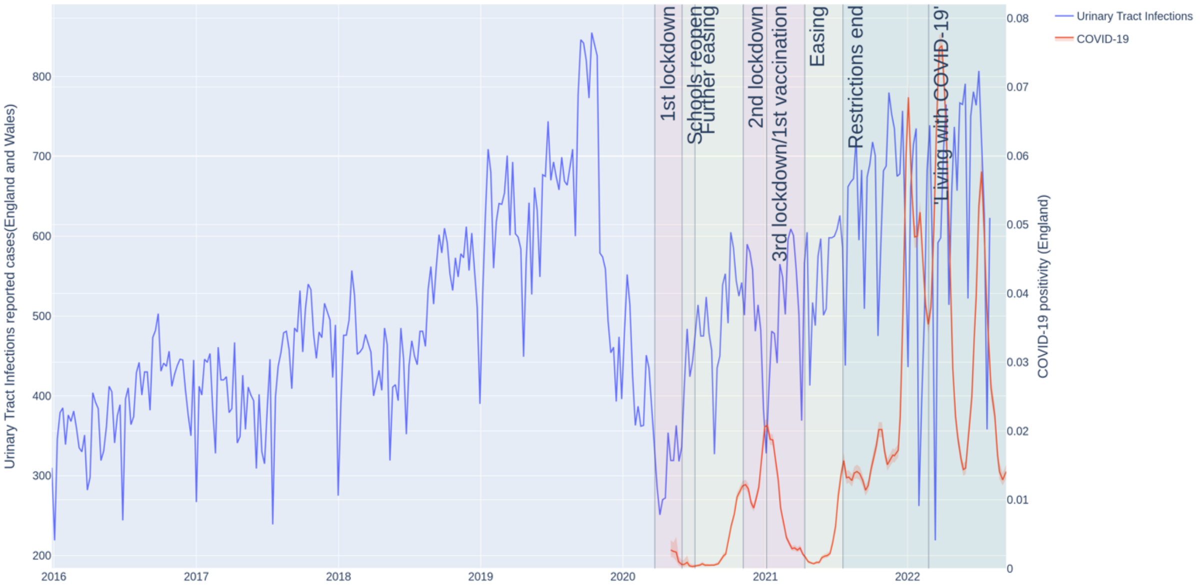


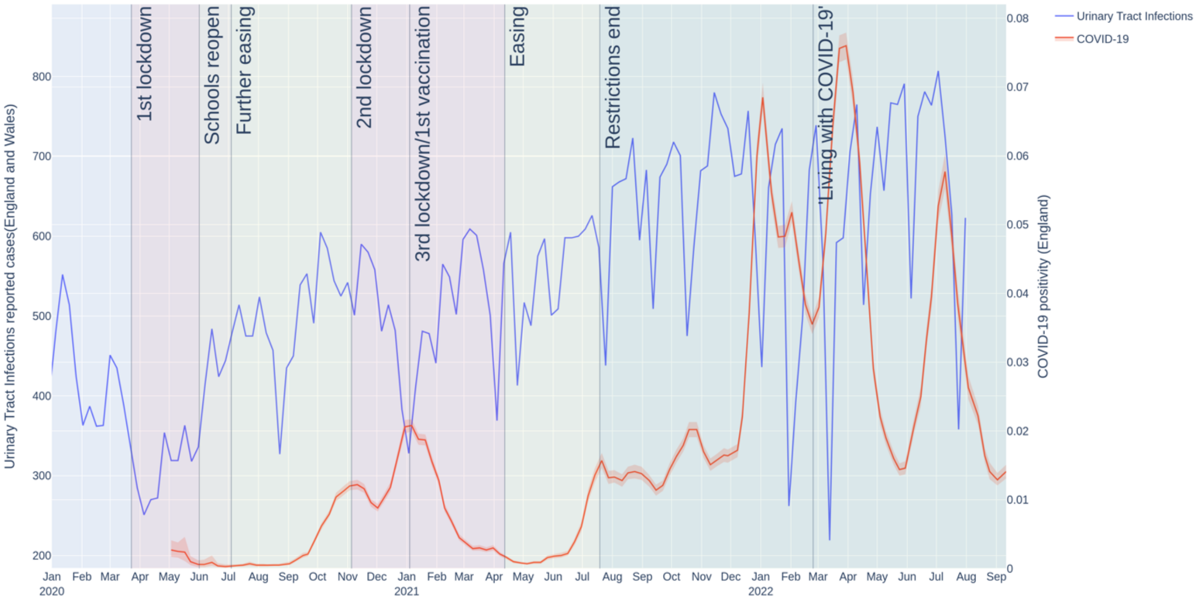


**Suppl. Figure 15. Urinary tract infection case numbers during the COVID-19 pandemic in England.** Weekly case numbers (Urinary tract infections: left y-axis, blue line, COVID-19: right y-axis, orange line) starting from 1st January 2016 (top graph) or 30th December 2019 (bottom graph).

**Suppl. Figure 16**

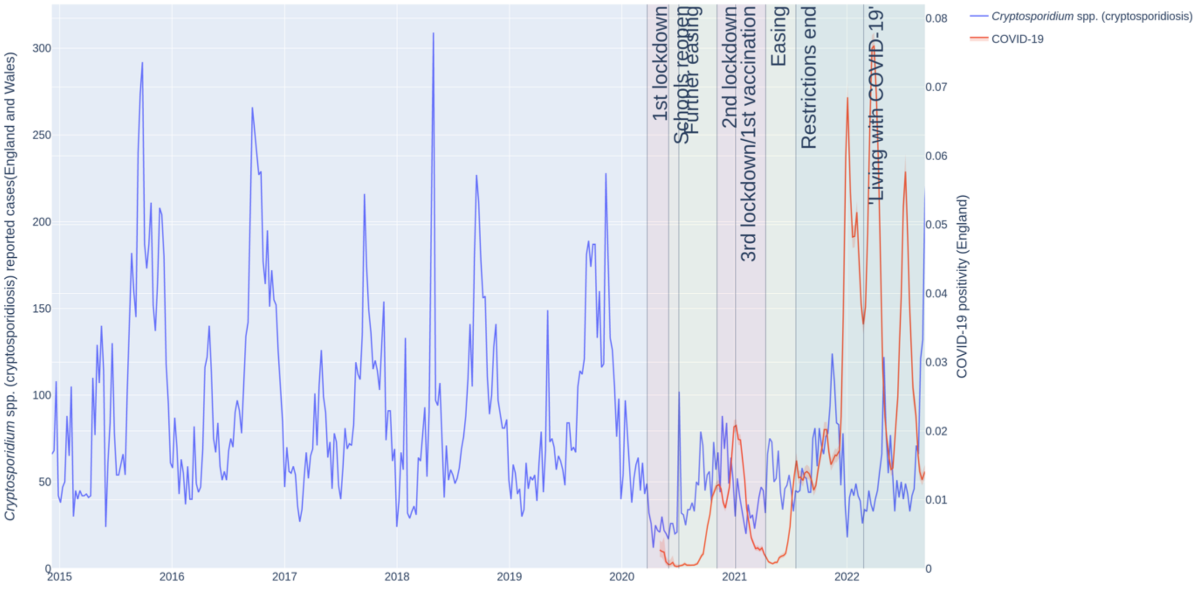


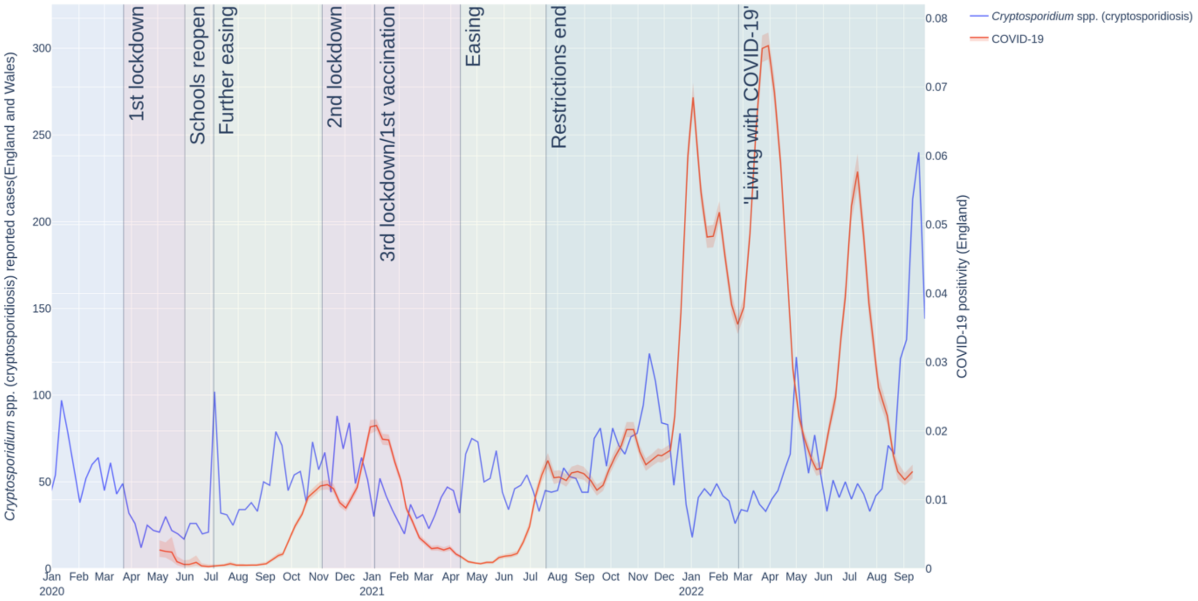


**Suppl. Figure 16. Cryptosporidiosis (*Cryptosporidium ssp.*) case numbers during the COVID-19 pandemic in England.** Weekly case numbers (*Cryptosporidium ssp.*: left y-axis, blue line, COVID-19: right y-axis, orange line) starting from 1st January 2015 (top graph) or 30th December 2019 (bottom graph).

**Suppl. Figure 17**

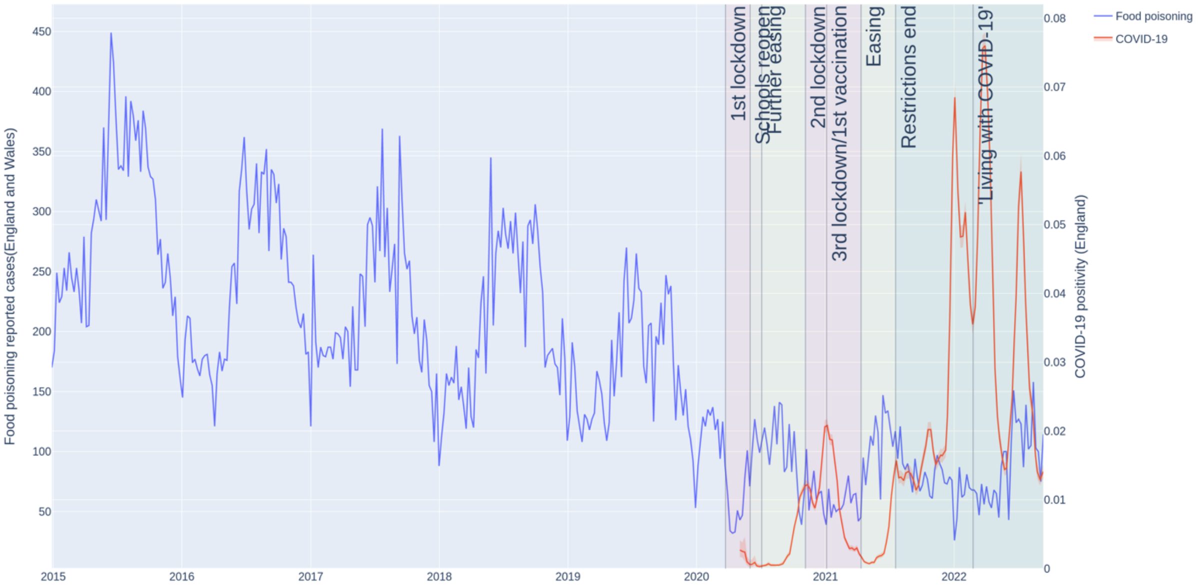


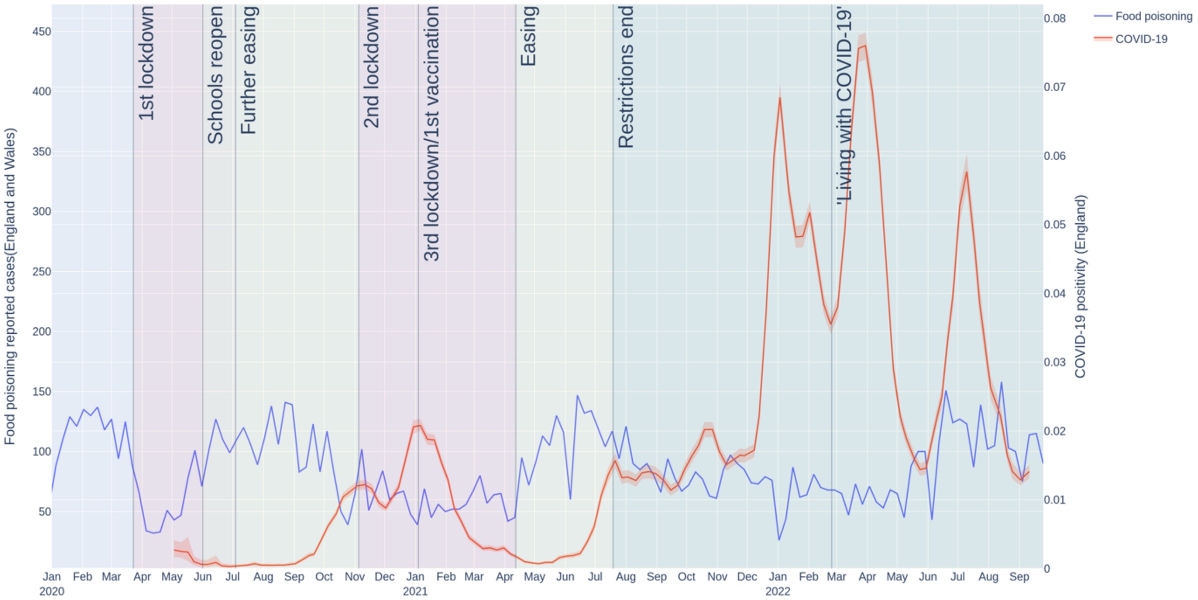


**Suppl. Figure 17. Foodborne illness (Food poisoning) case numbers during the COVID-19 pandemic in England.** Weekly case numbers (Food poisoning: left y-axis, blue line, COVID-19: right y-axis, orange line) starting from 1st January 2015 (top graph) or 30th December 2019 (bottom graph).

**Suppl. Figure 18**

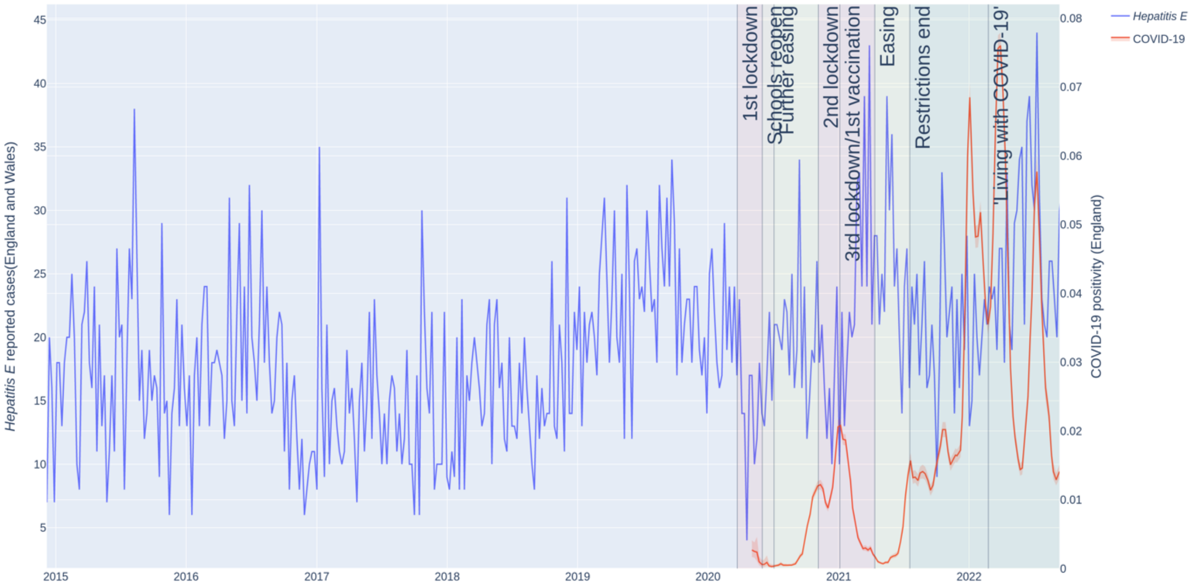


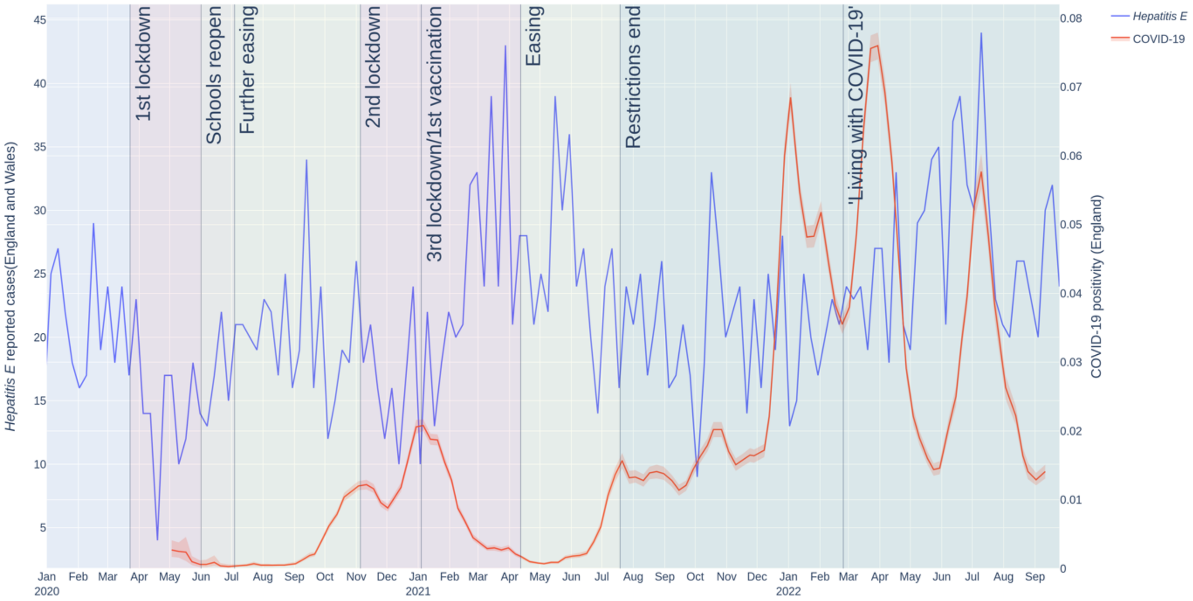


**Suppl. Figure 18. Hepatitis E case numbers during the COVID-19 pandemic in England.** Weekly case numbers (hepatitis E: left y-axis, blue line, COVID-19: right y-axis, orange line) starting from 1st January 2015 (top graph) or 30th December 2019 (bottom graph).

**Suppl. Figure 19**

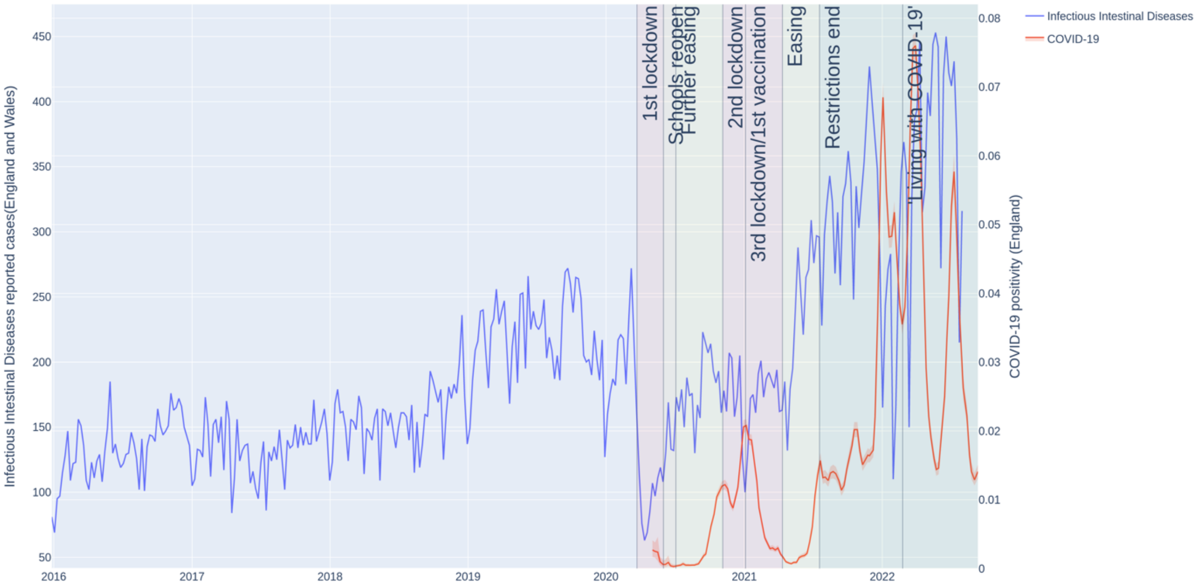


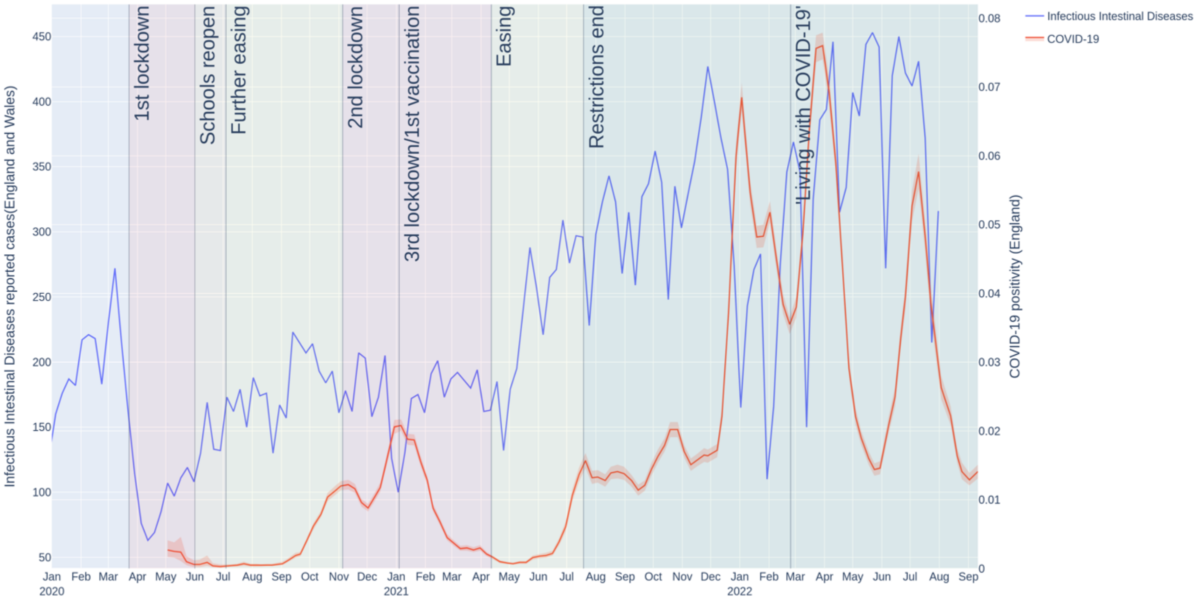


**Suppl. Figure 19. Infectious intestinal disease case numbers during the COVID-19 pandemic in England.** Weekly case numbers (Infectious intestinal disease: left y-axis, blue line, COVID-19: right y-axis, orange line) starting from 1st January 2016 (top graph) or 30th December 2019 (bottom graph).

**Suppl. Figure 20**

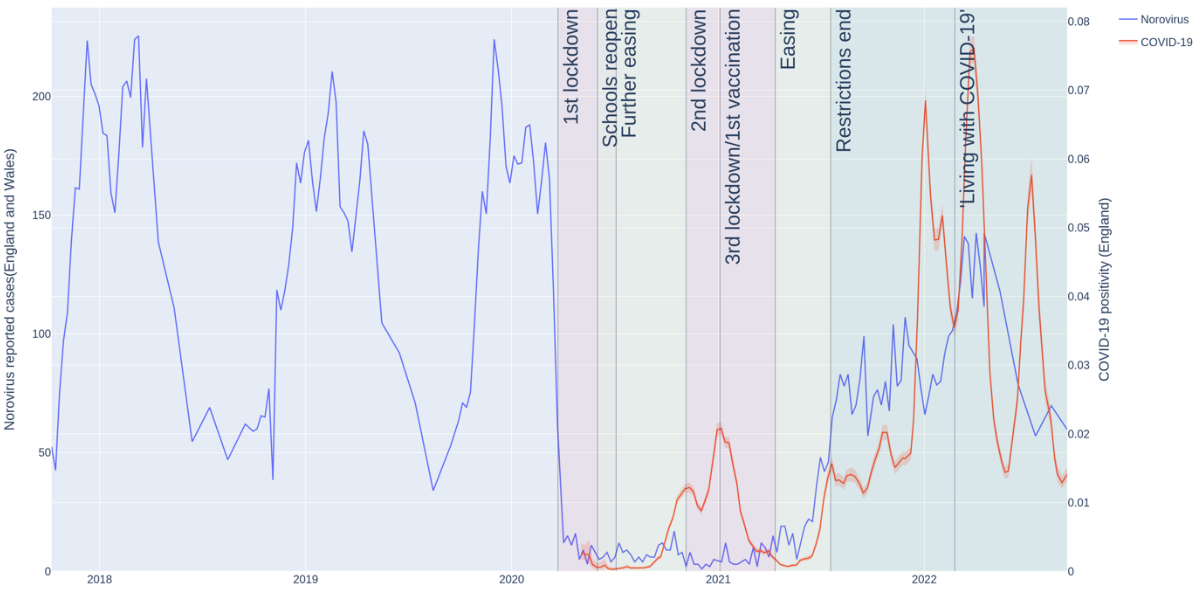


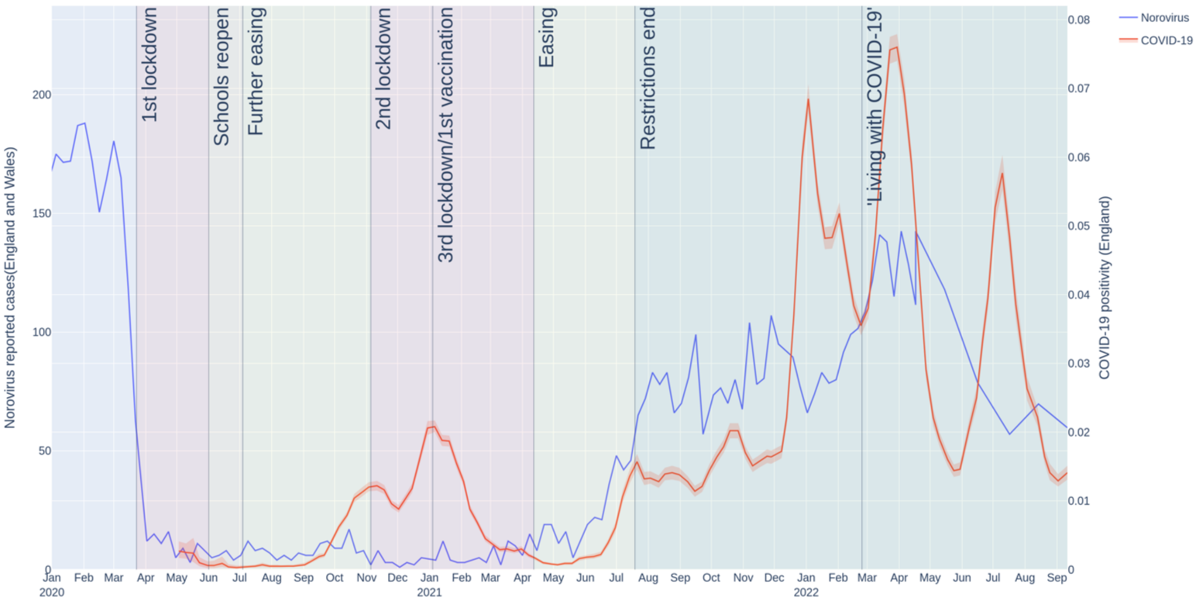


**Suppl. Figure 20. Norovirus case numbers during the COVID-19 pandemic in England.** Weekly case numbers (norovirus: left y-axis, blue line, COVID-19: right y-axis, orange line) starting from 2018 (top graph) or 30th December 2019 (bottom graph).

**Suppl. Figure 21**

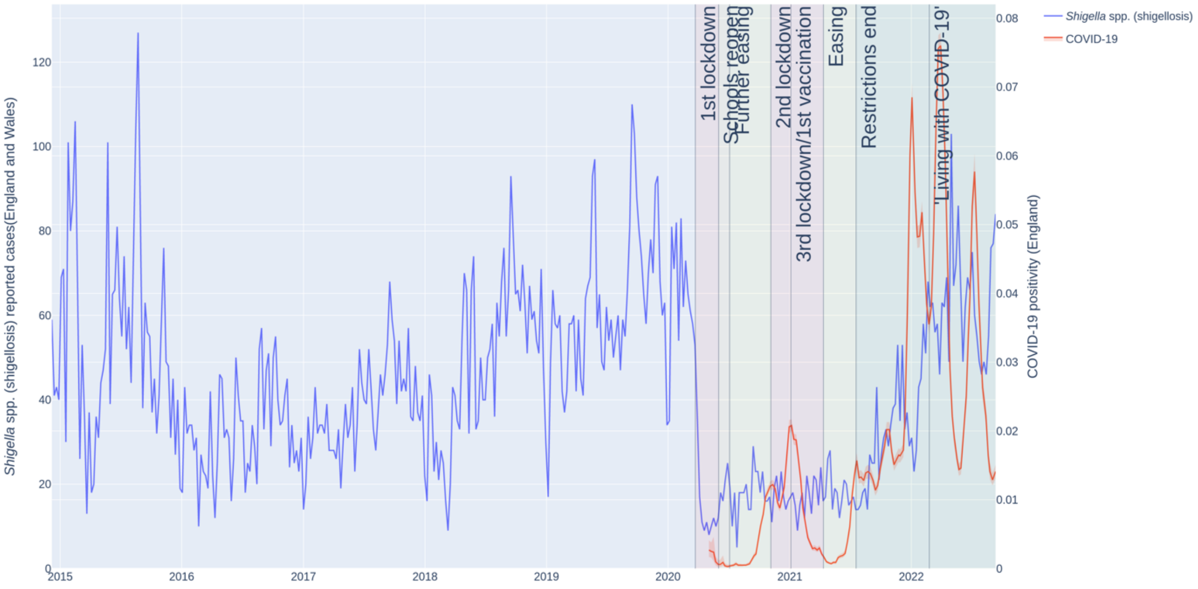


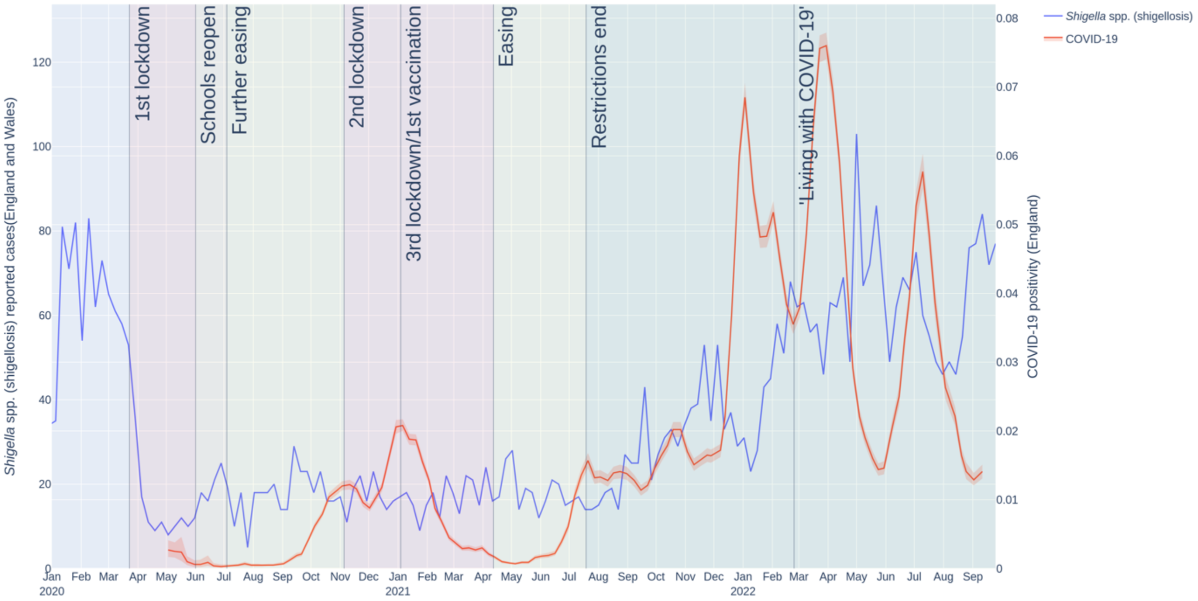


**Suppl. Figure 21. Shigellosis (*Shigella ssp.*) case numbers during the COVID-19 pandemic in England.** Weekly case numbers (*Shigella ssp.*: left y-axis, blue line, COVID-19: right y-axis, orange line) starting from 1st January 2015 (top graph) or 30th December 2019 (bottom graph).

**Suppl. Figure 22**

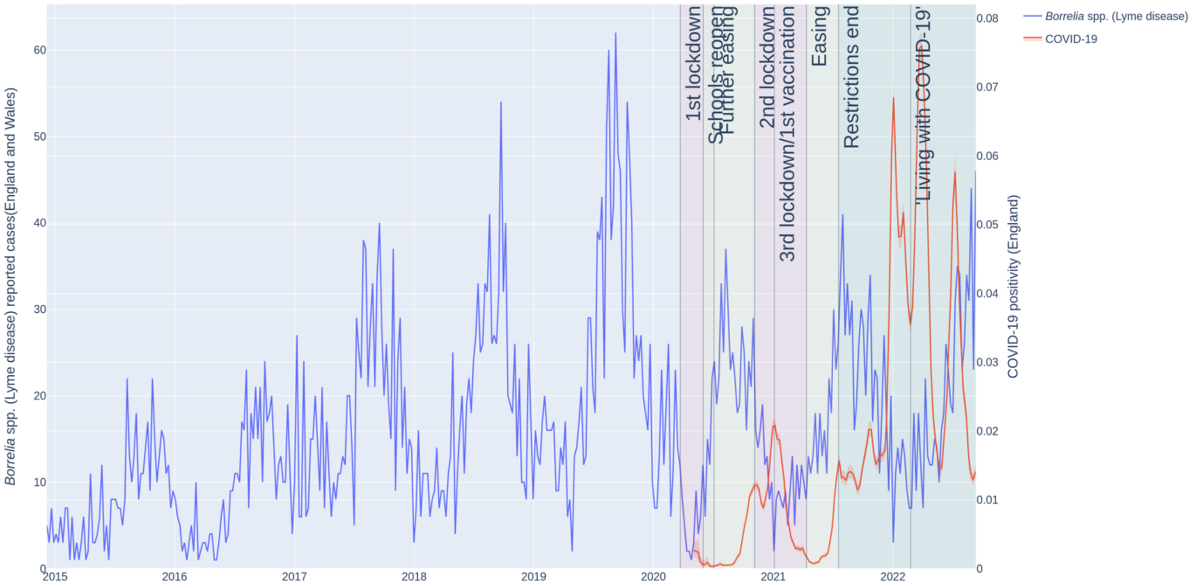


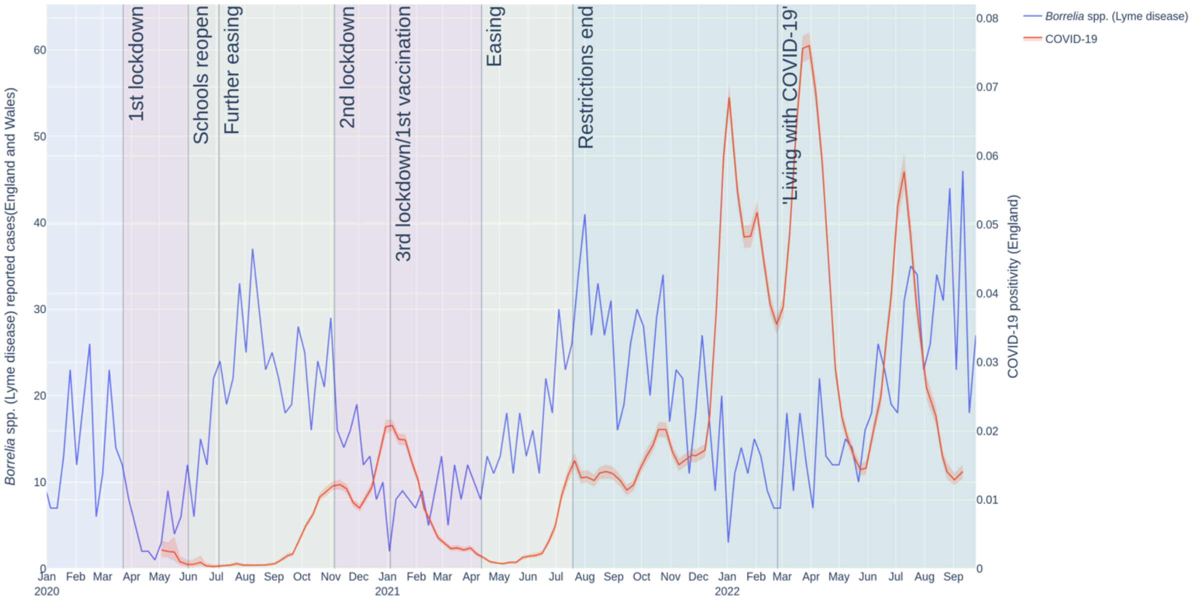


**Suppl. Figure 22. Lyme disease (*Borrelia ssp.*) case numbers during the COVID-19 pandemic in England.** Weekly case numbers (*Borrelia ssp.*: left y-axis, blue line, COVID-19: right y-axis, orange line) starting from 1st January 2015 (top graph) or 30th December 2019 (bottom graph).

**Suppl. Table 1.** Infectious diseases covered in this study and their anticipated modes of transmission.

| **Mode of transmission** | **Infectious diseases** |
|---|---|
| Airborne/droplet | Chickenpox, influenza-like-illness, measles, mumps, rubella, pneumococcal disease, scarlet fever, strep throat, tuberculosis, whooping cough |
| Blood-borne | Hepatitis C |
| Direct contact | Herpes simplex virus, methicillin resistant Staphylococcus aureus, skin and subcutaneous tissue infections, urinary tract infections |
| Faecal-oral | Cryptosporidiosis, foodborne illness, hepatitis E, infectious intestinal diseases, norovirus, shigellosis |
| Vector | Lyme disease |

**Suppl. Table 2.** Timing of COVID-19 protection measures in England.

| Year | Date | Week/Quarter | Prevention Measure | Guidelines |
|---|---|---|---|---|
| 2020 | 23rd March | 13/1 | First lockdown | - Confinement to household with exception for reasons deemed essential such as food or medicine.<br>- 2-meter social distancing.<br>- Mandatory wearing of face covering.<br>- Closure of schools and non-essential businesses.<br>- Ban of public gatherings.<br>- Cease of foreign travel.<br>- Rigorous hygiene measures including regular hand washing, use of hand sanitiser and frequent disinfection of surfaces. |
| | 1st June | 23/2 | Schools reopen | Phased reopening of nurseries, primary and secondary schools. |
| | 4th July | 27/3 | Further easing | Reopening of all non-essential business, such as restaurants, hairdressers, leisure facilities and tourist attractions. |
| | 5th November | 45/4 | Second lockdown | Schools and universities remained open. |
| 2021 | 4th/5th Jan | 1/1 | First vaccination/Third lockdown | Phased administration of the COVID-19 vaccine. |
| | 12th April | 15/2 | Easing | Reopening of all non-essential business. |
| | 19th July | 29/3 | Restrictions end | All restrictions lifted including removal of social distancing and reopening of existing closed sectors such as nightclubs. |
| 2022 | 24th February | 8/1 | ‘Living with COVID-19’ strategy | Legal requirement to self-isolate and wear face coverings removed. |
